# Identifying anaphylaxis using weakly-supervised prediction models and natural language processing

**DOI:** 10.64898/2026.06.09.26355005

**Authors:** Brian D Williamson, David J Cronkite, Onchee Yu, Arvind Ramaprasan, Sharon Fuller, Jennifer Covey, Erika Kiniry, Daniel Park, Robert Winter, Jill Whitaker, Michael F. McLemore, Saranrat Wittayanukorn, Danijela Stojanovic, Yueqin Zhao, Sarah Dutcher, David S Carrell, Lisa A Jackson, Jennifer C Nelson, Joshua C Smith

## Abstract

**Objectives:** Scalable computable phenotyping algorithms are critical for conducting high-throughput disease-outcome research in large, distributed-data electronic health record (EHR) and claims data settings. We developed and evaluated a claims- and EHR-based computable phenotyping algorithm for anaphylaxis, a rare acute condition that is challenging to accurately identify using claims data alone.

**Materials and Methods:** Potential anaphylaxis events came from two healthcare systems (Kaiser Permanente Washington [KPWA] and Vanderbilt University Medical Center [VUMC]). We engineered features from clinical text using automated natural language processing (NLP) methods. We then developed a phenotyping algorithm using four NLP- and diagnosis code-based silver labels (proxies for the gold-standard labels). Gold-standard abstracted outcomes were used to evaluate algorithm performance.

**Results:** The largest area under the receiver operating characteristic curve (AUC) was 0.931 for an NLP-based silver-label model at KPWA. Depending on the model and healthcare system site, positive predictive value (PPV) and sensitivity at the threshold of predicted probability that maximized F1 score ranged from 0.52 to 0.77 (PPV) and 0.78 to 1 (sensitivity).

**Discussion:** NLP-based silver-label models had large AUC at KPWA but not at VUMC. This may be because clinical text at KPWA is only available for outpatient encounters and secure messaging. High sensitivity for identifying anaphylaxis can be obtained using our best-performing models.

**Conclusion:** The best-performing models had better PPV and sensitivity tradeoffs than prior bespoke anaphylaxis models with costly, manually curated features. The simplicity of the approach compared to traditional phenotyping methods allows it to be deployed easily at multiple health care systems.

## Background and significance

Anaphylaxis, a rare but severe allergic reaction with rapid onset ^1^, is an important outcome for pharmacosurveillance, including vaccine safety studies. It can be difficult to define anaphylaxis algorithmically ^2^, creating a challenge for accurate identification of anaphylaxis using observational medical claims and electronic health records (EHR) data ^3^. Anaphylaxis diagnosis codes alone have been shown to be inadequate for case identification, with positive predictive value (PPV) ranging from 0.38 to 0.72 and sensitivity ranging from 0.6 to 0.9 ^4–7^; additional features are often required but have not yielded substantial gains in PPV or sensitivity ^3,8–10^. Recent work using features derived from natural language processing (NLP) of clinical notes has shown improvements in PPV for anaphylaxis and other challenging outcomes ^3,10–13^.

Computable phenotyping algorithms utilize EHR data, including NLP of clinical notes, and structured medical claims data to better identify patients with specific health outcomes of interest ^3^. These algorithms have traditionally relied on gold-standard training data (based on time- and expert-intensive manual chart review) and laborious manual engineering of candidate predictor variables derived from both structured data and unstructured text. The high time and cost requirements of this traditional approach limit not only the size of training datasets and the number of candidate predictor variables that can be considered during modeling, but also how many algorithms can be developed given a fixed amount of resources. The general-purpose PheNorm algorithm ^14^ provides a more scalable approach to computable phenotyping. PheNorm is a weakly-supervised method ^15^: it uses “silver label” proxies for gold-standard outcomes in model training. It can easily be combined with methods to automate NLP feature extraction, further reducing development time ^16,17^. While originally designed for common chronic conditions, PheNorm has recently been successfully applied to an acute condition, COVID-19 disease ^18^.

In this article, we use PheNorm to create a computable algorithm to identify patients with anaphylaxis using claims and EHR data. We develop two algorithms, using data from Kaiser Permanente Washington and Vanderbilt University Medical Center in a distributed-data setting, i.e., where source data remain behind health care system firewalls ^19,20^. These algorithms aim to fulfill a key goal of the US Food and Drug Administration (FDA) Sentinel Initiative ^21–24^ and the US Centers for Disease Control and Prevention (CDC) Vaccine Safety Datalink (VSD) ^25–27^ to rapidly and accurately identify acute health outcomes of interest for safety surveillance studies.

## Objective

We develop computable phenotyping models to identify patients with anaphylaxis. We evaluate performance in two different healthcare settings. We first consider internal model performance, where a model is evaluated in the same system it was developed in. Second, we consider external model performance, where a model is evaluated in the other system. Finally, in one health system we evaluate performance among participants known to have had a vaccination 0-2 days prior to the index encounter (vaccine-proximal encounters) and those known not to have had a vaccination during this time period, an important first step towards using computable phenotyping algorithms for vaccine safety surveillance.

## Methods

### Settings and study cohorts

Eligible patients received care at one of two healthcare systems: Vanderbilt University Medical Center (VUMC) and Kaiser Permanente Washington (KPWA). VUMC is an academic medical center delivering emergency, inpatient, and outpatient care through over 3 million patient visits per year in the southern United States. KPWA is a health maintenance organization providing integrated care to over 660,000 patients through 25 outpatient clinics in Washington state; inpatient and emergency care are externally contracted.

Patient encounters were included in the study if they met one of the eligibility criteria defined in Table 1. All VUMC data came from the VUMC EHR (includes billing codes and other diagnosis codes that were not used for billing); KPWA data came from the KPWA outpatient EHR (including urgent care and secure messaging) supplemented by structured medical claims data for inpatient, emergency, and outpatient care provided to KPWA enrollees by external providers. Clinical notes at KPWA are only available from the KPWA EHR. This activity was conducted under the authority of the FDA Sentinel Initiative in support of FDA medical product safety surveillance and under the authority of the Vaccine Safety Datalink for the CDC. Work conducted under FDA authority was a public health surveillance activity and, accordingly, was not subject to Institutional Review Board (IRB) oversight ^28–30^. For work conducted under CDC authority, the KPWA IRB determined this to be research and IRB approval was obtained.

**Table 1:**
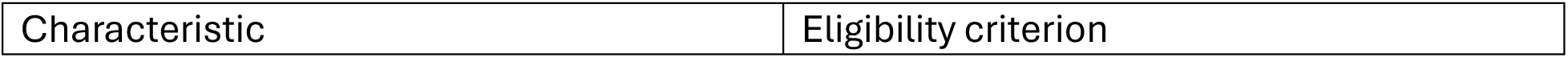
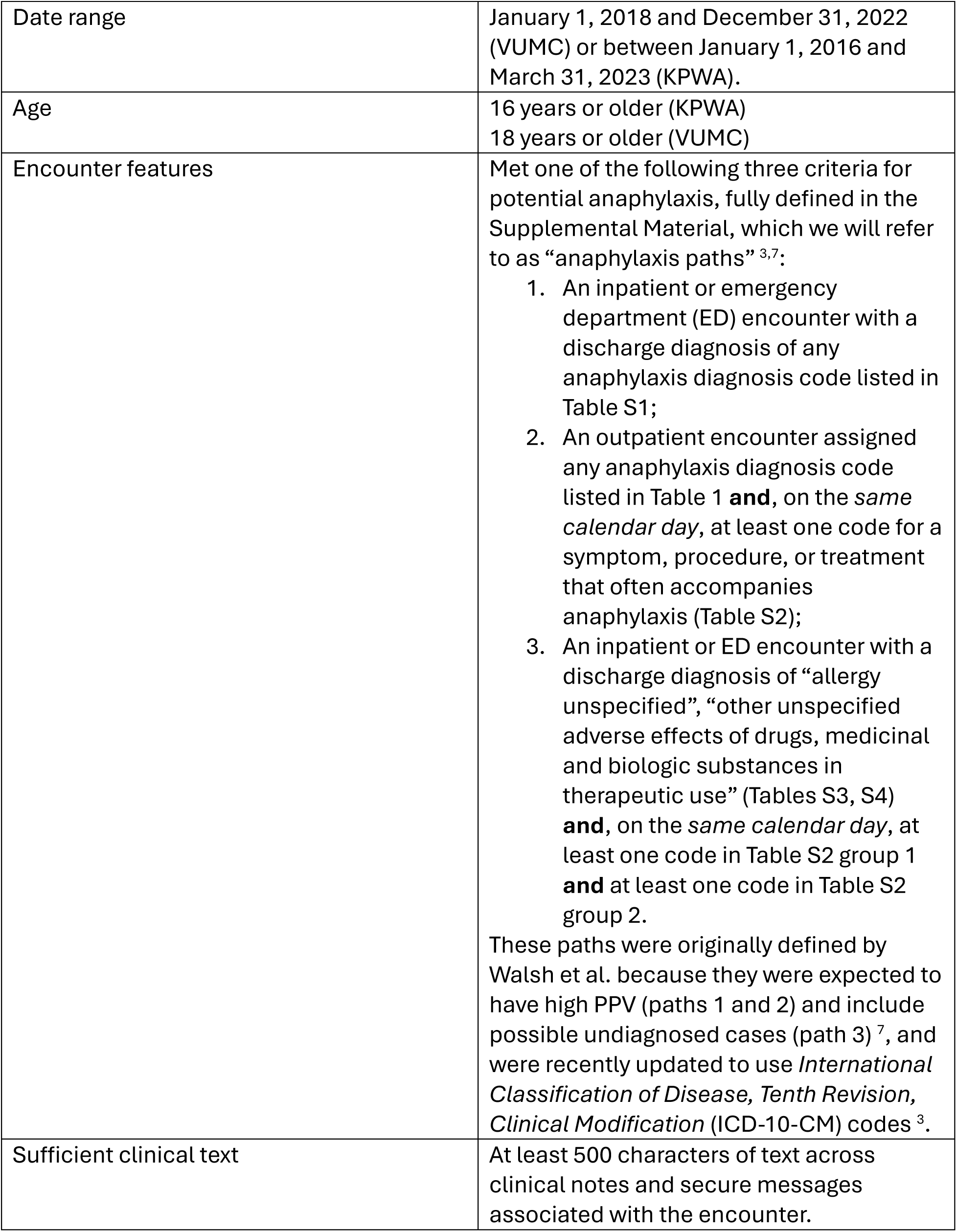
Eligibility criteria for medical encounters.

### Data catchment period for potential anaphylaxis episodes

A fixed data-catchment window is required to operationalize true outcomes, silver-standard labels (defined below), and features. Anaphylaxis is an acute condition, which requires a narrow window ^18^. The index encounter for a potential anaphylaxis episode corresponds to the first encounter in any 60-day period meeting one of the three paths defined above. The catchment period begins two days prior to the qualifying encounter and ends on the 7^th^ day following the qualifying encounter. For inpatient or ED encounters this is day 7 after discharge. After the episode ends, the patient could have another qualifying index encounter after a 60-day washout period. All potential anaphylaxis episodes for a given patient were included; a small proportion of patients had multiple episodes.

We further required the encounter to have at least 500 characters of text in clinical notes (including secure messaging) associated with the encounter during the data catchment period to ensure that there was a minimal amount of text to generate meaningful NLP features. All notes within each catchment window were processed using the MetaMap ^31,32^ NLP tool to identify mentions of concepts represented in the Unified Medical Language System (UMLS) ^33^. NLP-extracted concepts were represented using UMLS concept unique identifiers (CUIs).

### Silver-standard labels

Training a prediction model using gold-standard outcomes, which requires extensive manual medical record review – a costly and time-consuming task, limiting the number of reviewed records. In contrast, PheNorm is *weakly-supervised*: it uses easily-operationalized *silver-standard* labels that are hypothesized to be correlated with the true (gold-standard) outcomes for training. This reserves limited gold-standard outcomes for model evaluation and allows the PheNorm algorithm to be trained on many more observations than those in the gold-standard subset. We created four silver labels based on structured and NLP data, following the approach of the original PheNorm developers and the silver labels successfully used to identify COVID-19 ^14,18^:

1. Anaphylaxis diagnosis codes from claims and EHR: the count of unique encounters in the episode coded with any of the ICD-10-CM diagnosis codes for anaphylaxis listed in Table S1;
2. Anaphylaxis mentions: NLP-based counts of all mentions of anaphylaxis in chart notes in the episode, identified by the case-insensitive regular expression “\banaph\w*” (i.e., words that start with the string “anaph” and are followed by zero or more characters);
3. Anaphylaxis CUIs: NLP-based counts of notes containing one or more strings tagged with any UMLS CUI representing anaphylaxis and/or anaphylaxis treatment. The CUIs used were C4316895 (anaphylactic shock), C0685898 (anaphylactic shock due to adverse food reaction), C0340865 (anaphylactoid reaction), C0002792 (anaphylaxis), C0854649 (anaphylaxis treatment);
4. Anaphylaxis or epinephrine mentions: NLP-based counts of all mentions of anaphylaxis or epinephrine in chart notes during the episode, identified by either of the following two case-insensitive regular expressions: “\banaph\w*” or “\bepine\w*” (i.e., words that start with the string “epine” and are followed by zero or more characters).

### Feature engineering

We used both structured data features from claims and EHR and features extracted from clinical text using NLP in our PheNorm prediction models. Structured data features include: a binary indicator of vaccine administration within 0—2 days prior to the index encounter (at KPWA only; we refer to this as “vaccine proximal”); a binary indicator of evidence in structured data that the patient received a procedure for administration of antihistamine during the encounter; a binary indicator of evidence in structured data that the patient received a procedure for administration of epinephrine during the encounter; age at index date (years); sex (as captured in the EHR); and race and Hispanic ethnicity (as captured in the EHR). Vaccine status was only captured at KPWA due to its participation in the Vaccine Safety Datalink.

We selected NLP features (anaphylaxis-relevant UMLS concepts) using the automated feature extraction for phenotyping (AFEP) method ^16^. Briefly, we used MetaMap to identify UMLS concepts with relevant semantic types that appeared in clinical knowledge-base articles on anaphylaxis (MedlinePlus, Medscape, Merck, Mayo Clinic, and Wikipedia). Concepts found in a majority (at least three of five) of the articles are more likely to be relevant to anaphylaxis. Features based on these concepts are the raw counts of each concept appearing in a patient’s notes during the data catchment window. To address potential omissions due to text variability, we also counted instances of any AFEP-identified concepts’ associated child concepts (based on vocabulary hierarchy) as if they were mentions of the parent concept. We utilized the default MetaMap scoring options and restricted CUIs to the RxNorm and MedDRA ^34^ vocabularies when processing both the knowledge-base articles and clinical notes.

### Gold-standard outcomes

Gold-standard outcomes were defined using chart abstraction to determine if an event was likely anaphylaxis based on the National Institute of Allergy and Infectious Diseases/Food Allergy and Anaphylaxis Network (NIAID/FAAN) criteria for anaphylaxis ^35^. A medical record abstraction form (see Supplementary Material) was developed at VUMC and used by trained abstractors on records at VUMC and KPWA. Using the form, abstractors collected data from the medical record on timing of onset, exposure to likely or known allergens, and symptoms/organ system involvement. The date range for medical records included in chart abstraction is the data catchment period defined above. These abstracted data were then used with a pre-specified algorithm to make a case determination based on the NIAID/FAAN criteria for anaphylaxis. Medical record abstractions were performed at VUMC using emergency room, inpatient, and outpatient records (including secure messaging). At KPWA, because medical records from emergency room and inpatient encounters for KPWA members are not available for processing using NLP, only urgent care and outpatient records (including secure messaging) were used for the medical record abstraction (data from emergency and inpatient encounters were used to create silver labels and features). These visits could either be the index visit or an outpatient visit (or secure message) following an index visit in an emergency or inpatient setting.

### Gold-standard sample

The VUMC gold-standard sample is a random sample stratified by anaphylaxis path (Table 1). Thirty charts were initially abstracted by two nurse informaticist abstractors to assess inter-rater reliability (Cohen’s kappa 0.67); subsequent abstractions were performed by only one abstractor. The final gold-standard sample is slightly over-sampled for Paths 2 and 3 to provide sufficient training data from these paths, which we anticipated to be more challenging for the algorithm to classify.

In the KPWA gold-standard stratified random sample we over-sampled Paths 2 and 3 and vaccine-proximal encounters. All 40 episodes with vaccine-proximal index encounters were included in the sample. A further 105 non-vaccine-proximal encounters were then sampled stratified by anaphylaxis path defined above and whether the patient’s medical record number ended in an even or odd digit (this final stratum was created for administrative reasons because charts were reviewed at two separate times). The final strata, along with number eligible and selected per stratum and the post-stratification sampling weight ^36^, are provided in Table S5. A subset (n=50) of charts were abstracted by multiple abstractors (Cohen’s kappa 0.634); the remaining charts were abstracted by a single abstractor.

### Model development

We developed separate anaphylaxis identification models at VUMC and KPWA using the same sets of features from structured and unstructured data. We used a train/test split for model training (episodes not sampled for gold-standard labeling) and evaluation (on the gold-standard sample). We applied a site-specific filter, where if any CUIs had zero mention at one of the sites, they were dropped from the set of predictor variables for that site. We did not apply further dimension reduction, exclude negated mentions, or normalize CUI variables, following Smith et al. ^18^.

PheNorm uses weakly-supervised linear regression with corruption and the expectation-maximization (EM) algorithm ^37^ to create a predicted probability of anaphylaxis based on each silver label. First, for a given silver label *S*_*j*_, a corrupted and log-transformed value is created: 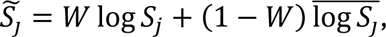 where *W* ∈ {0,1} is a binary variable with Pr(*W* = 1) = *r* (for tuning parameter *r* ∈ (0,1)) and 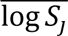 the mean value of log *S*_*j*_. This is used to estimate linear regression parameters 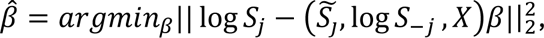 where *S*_−*j*_ are the remaining silver labels and *X* are the (NLP and clinical) features. The EM algorithm is then used to estimate parameters in a two-component Gaussian mixture model on the PheNorm score *Z* = (log *S*, *X*)*β̂*. The posterior probability from the EM algorithm is the silver label-based predicted probability.

We applied PheNorm with corruption rate *r* = 0.3 and training set multiplication factor 13, which are default values ^14^ that have worked well in another acute-outcome context ^18^, using all data without gold-standard outcome labels as training data. In addition to predicted probabilities from each silver label defined above, we also obtained predicted probabilities from the “aggregate” model, defined as the mean of the four silver label-specific probabilities.

### Model evaluation

We used standard prediction performance measures for binary outcome prediction settings to evaluate model performance, including area under the receiver operating characteristic curve (AUC), sensitivity, specificity, PPV, negative predictive value (NPV), F1 score, and F0.5 score. The F scores are the weighted harmonic mean of PPV and sensitivity; F1 gives each equal weight, while F0.5 gives higher weight to PPV. We computed these metrics based on predictions from the models developed above in each gold-standard sample. Because both samples were stratified random samples, we computed inverse-weighted versions of these prediction performance metrics to allow generalization back to the full populations; the sampling weights are provided in Table S5. We provide 95% confidence intervals (CIs) for AUC using the method of LeDell et al. ^38^. We consider AUC to represent the primary global assessment of model performance, because it does not rely on a threshold of predicted probability, but the other metrics provide important secondary performance assessments.

The code used for this analysis is available at https://github.com/kpwhri/Sentinel-Scalable-NLP/tree/master/Prediction-Modeling/Anaphylaxis. We used this code to analyze data at both KPWA and VUMC.

## Results

### Description of KPWA and VUMC samples

We present characteristics of both samples in Table 2. At VUMC, less than 1% of initial index encounters with a potential anaphylaxis episode were excluded due to having <500 characters of text across notes. There were 1283 eligible index encounters from 1221 patients. Of the eligible index encounters, 254 were sampled for gold-standard label ascertainment via chart review; 175 (68%) of these encounters were determined to be a true anaphylaxis event. At KPWA, approximately 40% of initial index encounters were excluded due to having <500 characters of text across notes. There were 1028 eligible index encounters from 979 patients. Of the eligible encounters, 145 were sampled for gold-label ascertainment; 58 (40%) of these encounters were determined to be a true anaphylaxis event. Most encounters were associated with 6 or fewer days with clinical notes.

**Table 2.** Characteristics of the study cohorts by study site.

| Characteristic | VUMC (N = 1283 encounters) |  | KPWA (N = 1028 encounters) |  |
| --- | --- | --- | --- | --- |
|  | Count | Percent | Count | Percent<sup>1</sup> |
| Number of encounters with gold-standard outcomes | 254 | 20% | 145 | 14% |
| Sex is female | 774 | 60% | 680 | 66% |
| Age group |  |  |  |  |
| 16—29 | 377 | 29% | 197 | 19% |
| 30—49 | 402 | 31% | 333 | 32% |
| 50—69 | 384 | 30% | 357 | 35% |
| 70+ | 120 | 9% | 141 | 14% |
| Race is White | 984 | 77% | 691 | 67% |
| Ethnicity is Hispanic | 64 | 5% | 54 | 5% |
| Anaphylaxis path<sup>2</sup> |  |  |  |  |
| 1 | 904 | 70% | 467 | 45% |
| 2 | 177 | 14% | 405 | 39% |
| 3 | 202 | 16% | 156 | 15% |
| Days with clinical notes |  |  |  |  |
| 1—3 days | 718 | 56% | 754 | 73% |
| 4—6 days | 225 | 18% | 234 | 23% |
| 7—9 days | 115 | 9% | 34 | 3% |
| 10+ days | 225 | 18% | 6 | 1% |

<sup>1</sup>Percentages may not add up to 100% due to rounding.
<sup>2</sup>Path is hierarchical: if an episode qualified through path 1, it was assigned path 1; if not, it was assessed for path 2 (and assigned to path 2 if it met the criteria), then path 3.

Applying AFEP to our five knowledge-base articles identified 159 clinical concepts associated with anaphylaxis that were used to operationalize NLP features (provided in the Supplementary Material Table S6). Of these 159 NLP features, 80 were MedDRA preferred term concepts with 622 associated child (lower-level term) concepts counted as instances of the parent. For example, any mentions of "Face angioedema" (C0743747) were counted as a mention for the included parent feature "Angioedema" (C0002994). Supplementary Data (Supplementary Material Section S2.2) contains the full list of child/parent mappings. From the full set of 159 NLP features, 144 were retained for use at KPWA and 153 at VUMC after screening out features with zero mentions.

### Primary global assessment of anaphylaxis model accuracy

We observed moderate but stable and similar AUC for both the KPWA and VUMC anaphylaxis diagnosis code PheNorm models (silver label 1) regardless of evaluation data site (Table 3; Figures S1—S4). The KPWA model had AUC 0.684 (95% CI [0.597, 0.771]) when evaluated on the KPWA gold-label data and 0.660 (0.584, 0.737) on the VUMC gold-label data. The VUMC model had AUC 0.749 (0.667, 0.831) on KPWA data and 0.664 (0.590, 0.739) on VUMC data. The diagnosis code-based models had the lowest AUC among silver labels on the KPWA data but the highest AUC on the VUMC data.

**Table 3.**
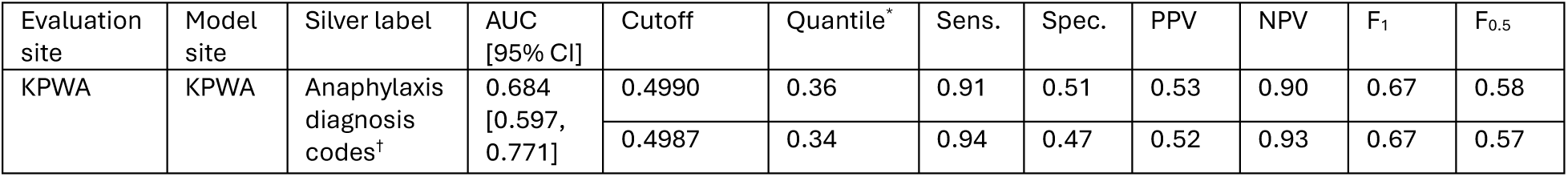
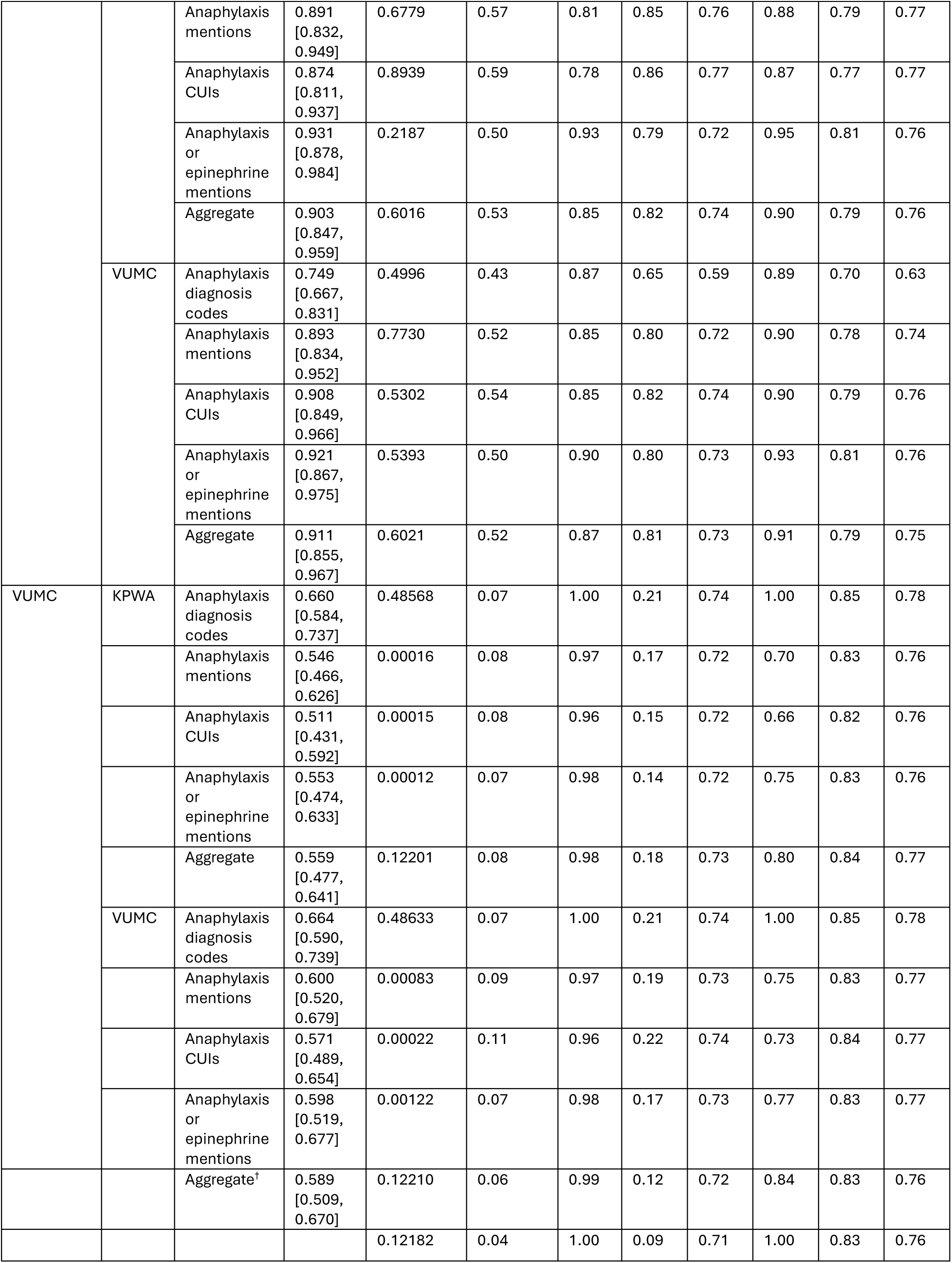
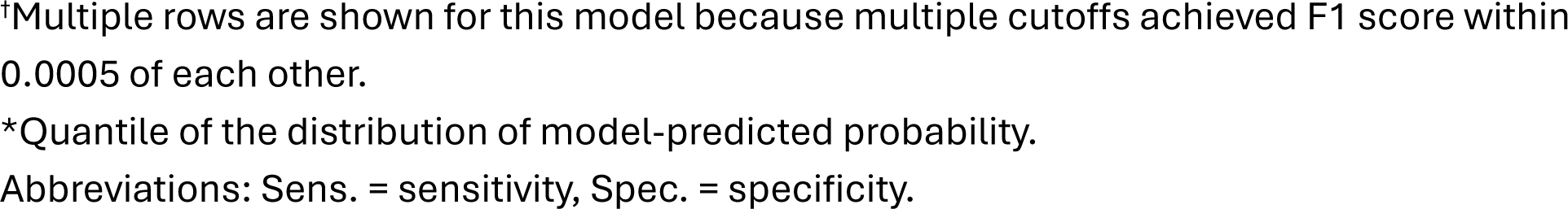
Performance of PheNorm models on KPWA and VUMC gold-standard episodes. For evaluation at KPWA, prediction performance of models based on each silver label evaluated on the 145 episodes with gold-standard labels at KPWA. For evaluation at VUMC, prediction performance on the 254 episodes with gold-standard labels at VUMC. Results include AUCs and 95% CIs and other performance metrics at the cutoff of predicted probability that maximized F_1_ score.

| Evaluation site | Model site | Silver label | AUC [95% CI] | Cutoff | Quantile* | Sens. | Spec. | PPV | NPV | $F_1$ | $F_{0.5}$ |
| --- | --- | --- | --- | --- | --- | --- | --- | --- | --- | --- | --- |
| KPWA | KPWA | Anaphylaxis diagnosis codes<sup>†</sup> | 0.684 [0.597, 0.771] | 0.4990 | 0.36 | 0.91 | 0.51 | 0.53 | 0.90 | 0.67 | 0.58 |
|  |  |  |  | 0.4987 | 0.34 | 0.94 | 0.47 | 0.52 | 0.93 | 0.67 | 0.57 |
|  |  | Anaphylaxis mentions | 0.891<br>[0.832, 0.949] | 0.6779 | 0.57 | 0.81 | 0.85 | 0.76 | 0.88 | 0.79 | 0.77 |
|  |  | Anaphylaxis CUIs | 0.874<br>[0.811, 0.937] | 0.8939 | 0.59 | 0.78 | 0.86 | 0.77 | 0.87 | 0.77 | 0.77 |
|  |  | Anaphylaxis or epinephrine mentions | 0.931<br>[0.878, 0.984] | 0.2187 | 0.50 | 0.93 | 0.79 | 0.72 | 0.95 | 0.81 | 0.76 |
|  |  | Aggregate | 0.903<br>[0.847, 0.959] | 0.6016 | 0.53 | 0.85 | 0.82 | 0.74 | 0.90 | 0.79 | 0.76 |
|  | VUMC | Anaphylaxis diagnosis codes | 0.749<br>[0.667, 0.831] | 0.4996 | 0.43 | 0.87 | 0.65 | 0.59 | 0.89 | 0.70 | 0.63 |
|  |  | Anaphylaxis mentions | 0.893<br>[0.834, 0.952] | 0.7730 | 0.52 | 0.85 | 0.80 | 0.72 | 0.90 | 0.78 | 0.74 |
|  |  | Anaphylaxis CUIs | 0.908<br>[0.849, 0.966] | 0.5302 | 0.54 | 0.85 | 0.82 | 0.74 | 0.90 | 0.79 | 0.76 |
|  |  | Anaphylaxis or epinephrine mentions | 0.921<br>[0.867, 0.975] | 0.5393 | 0.50 | 0.90 | 0.80 | 0.73 | 0.93 | 0.81 | 0.76 |
|  |  | Aggregate | 0.911<br>[0.855, 0.967] | 0.6021 | 0.52 | 0.87 | 0.81 | 0.73 | 0.91 | 0.79 | 0.75 |
| VUMC | KPWA | Anaphylaxis diagnosis codes | 0.660<br>[0.584, 0.737] | 0.48568 | 0.07 | 1.00 | 0.21 | 0.74 | 1.00 | 0.85 | 0.78 |
|  |  | Anaphylaxis mentions | 0.546<br>[0.466, 0.626] | 0.00016 | 0.08 | 0.97 | 0.17 | 0.72 | 0.70 | 0.83 | 0.76 |
|  |  | Anaphylaxis CUIs | 0.511<br>[0.431, 0.592] | 0.00015 | 0.08 | 0.96 | 0.15 | 0.72 | 0.66 | 0.82 | 0.76 |
|  |  | Anaphylaxis or epinephrine mentions | 0.553<br>[0.474, 0.633] | 0.00012 | 0.07 | 0.98 | 0.14 | 0.72 | 0.75 | 0.83 | 0.76 |
|  |  | Aggregate | 0.559<br>[0.477, 0.641] | 0.12201 | 0.08 | 0.98 | 0.18 | 0.73 | 0.80 | 0.84 | 0.77 |
|  | VUMC | Anaphylaxis diagnosis codes | 0.664<br>[0.590, 0.739] | 0.48633 | 0.07 | 1.00 | 0.21 | 0.74 | 1.00 | 0.85 | 0.78 |
|  |  | Anaphylaxis mentions | 0.600<br>[0.520, 0.679] | 0.00083 | 0.09 | 0.97 | 0.19 | 0.73 | 0.75 | 0.83 | 0.77 |
|  |  | Anaphylaxis CUIs | 0.571<br>[0.489, 0.654] | 0.00022 | 0.11 | 0.96 | 0.22 | 0.74 | 0.73 | 0.84 | 0.77 |
|  |  | Anaphylaxis or epinephrine mentions | 0.598<br>[0.519, 0.677] | 0.00122 | 0.07 | 0.98 | 0.17 | 0.73 | 0.77 | 0.83 | 0.77 |
|  |  | Aggregate<sup>†</sup> | 0.589<br>[0.509, 0.670] | 0.12210 | 0.06 | 0.99 | 0.12 | 0.72 | 0.84 | 0.83 | 0.76 |
|  |  |  |  | 0.12182 | 0.04 | 1.00 | 0.09 | 0.71 | 1.00 | 0.83 | 0.76 |

†Multiple rows are shown for this model because multiple cutoffs achieved F1 score within 0.0005 of each other.
\*Quantile of the distribution of model-predicted probability.
Abbreviations: Sens. = sensitivity, Spec. = specificity.

In contrast, we observed highly heterogeneous performance NLP-based silver label models both within a given evaluation dataset and across datasets. These models had large AUC when evaluated on the KPWA data (Table 3): at KPWA, the smallest AUC was 0.874 (0.811, 0.937) for the anaphylaxis CUIs model (silver label 3), while the largest AUC was 0.931 (0.878, 0.984) for the anaphylaxis or epinephrine mentions model (silver label 4). Performance was much worse when evaluated on the VUMC data (Table 3): the smallest AUC was 0.511 (0.431, 0.592) for the KPWA anaphylaxis CUIs model, while the largest AUC was 0.600 (0.520, 0.679) for the VUMC anaphylaxis mentions model (silver label 2).

### Secondary assessments of anaphylaxis model performance

PheNorm models can predict anaphylaxis with high sensitivity and reasonable tradeoffs with PPV. In Table 3, we present sensitivity, specificity, PPV, NPV, F1, and F0.5 at the cutoff of model-predicted probability of anaphylaxis that maximized F1 score (balances sensitivity and PPV equally). The KPWA anaphylaxis or epinephrine mentions model (silver label 4) evaluated at KPWA had sensitivity 0.93 and PPV 0.72 at the cutoff that maximized F1 score (at 0.81), while the aggregate model had sensitivity 0.85 and PPV 0.74 at the cutoff that maximized F1 score (0.79; Table 3). We show all metrics at all cutoffs for the KPWA aggregate model evaluated at KPWA in Figure 1. For example, sensitivity >0.9 could be achieved with PPV approximately 0.65. Similar tradeoffs can be observed for VUMC models evaluated on VUMC data (Table 3, Figure 2), though sensitivity tended to be higher for these models than the KPWA models evaluated on KPWA data.

**Figure 1:**
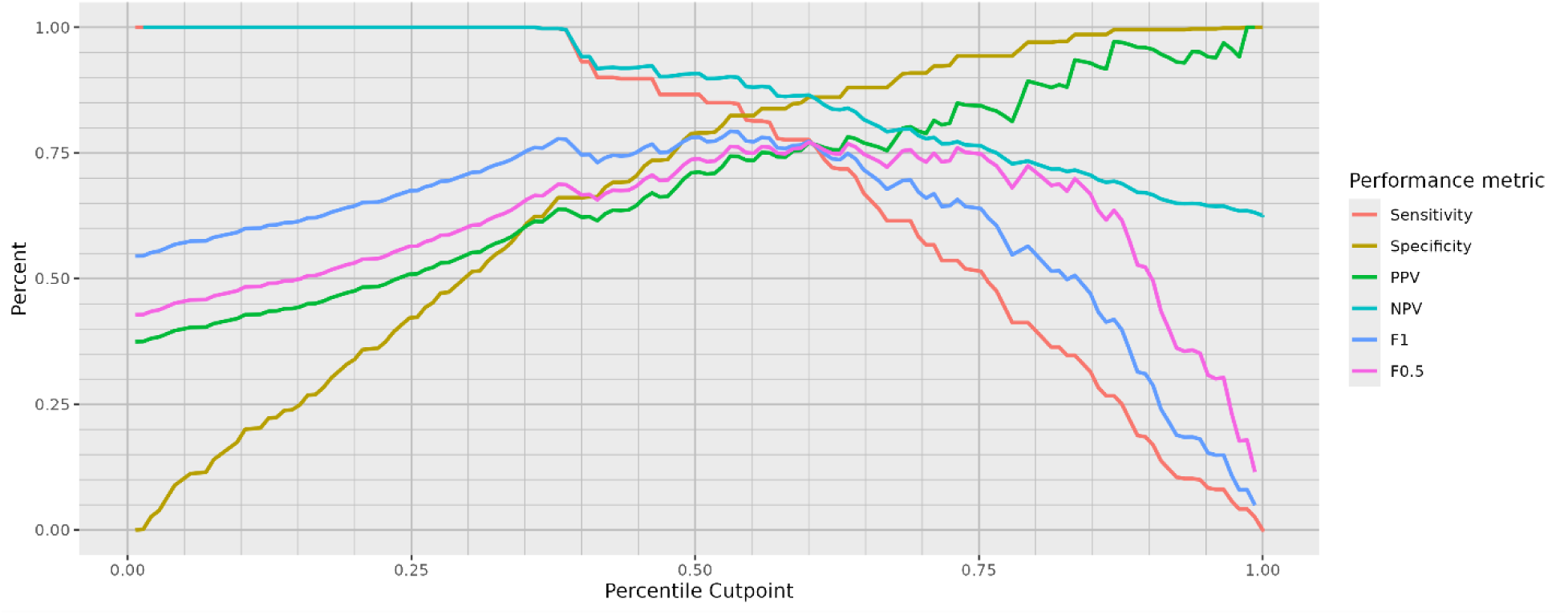
Model performance metrics at different cutpoints of predicted risk for the aggregate model trained at KPWA (using 883 episodes without gold-standard labels) and evaluated on 145 episodes with gold-standard labels from KPWA. The horizontal axis is the quantile of the distribution of model-predicted probability.

**Figure 2:**
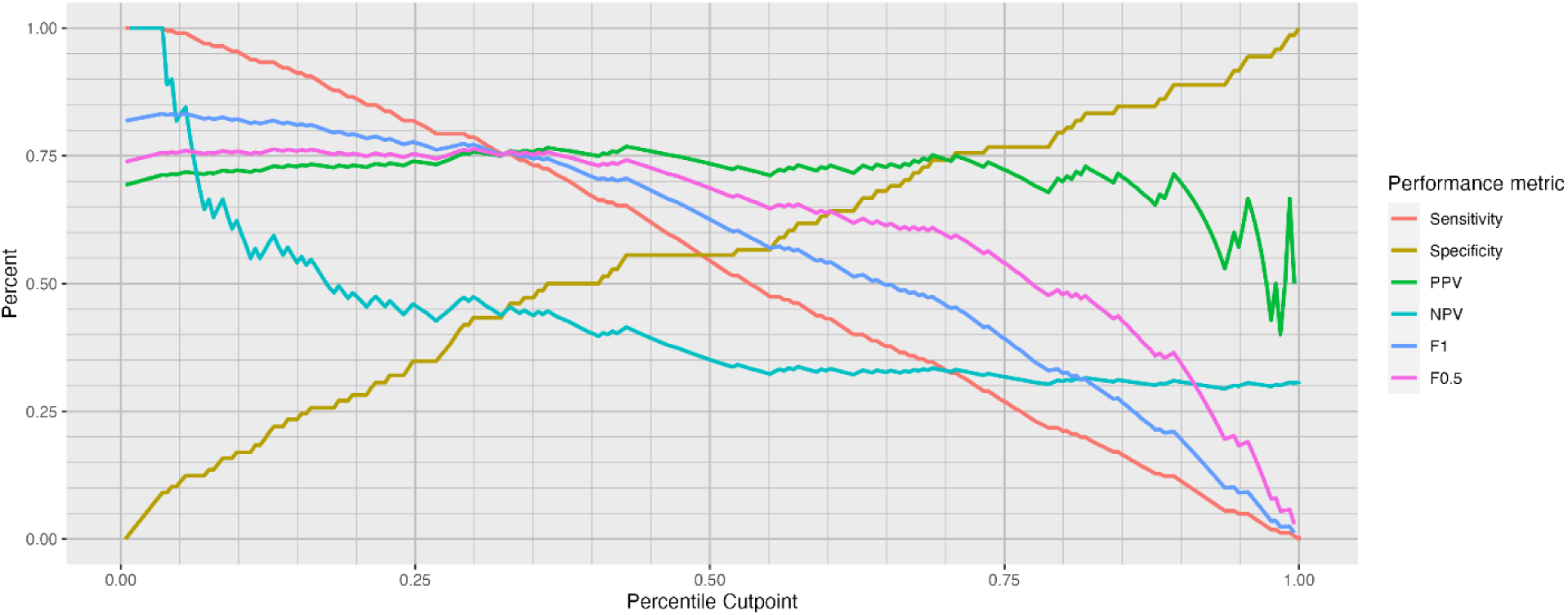
Model performance metrics at different cutpoints of predicted risk for the aggregate model trained at VUMC (using 1059 episodes without gold-standard labels) and evaluated on 254 episodes with gold-standard labels from VUMC. The horizontal axis is the quantile of the distribution of model-predicted probability.

We also considered cross-site performance evaluation. The VUMC model with the highest sensitivity on the KPWA data was the anaphylaxis or epinephrine mentions model (silver label 4), with sensitivity 0.9 and moderate PPV (0.73) at the cutoff that maximized F1 (Table 2). For the VUMC aggregate model on the KPWA data, the sensitivity and PPV at the cutoff that maximized F1 were 0.87 and 0.73, respectively (Table 3). Different tradeoffs could be made for either KPWA or VUMC models (Figures 3 and 4). Sensitivity tended to be higher for the KPWA models evaluated at VUMC than for the VUMC models evaluated at KPWA.

**Figure 3:**
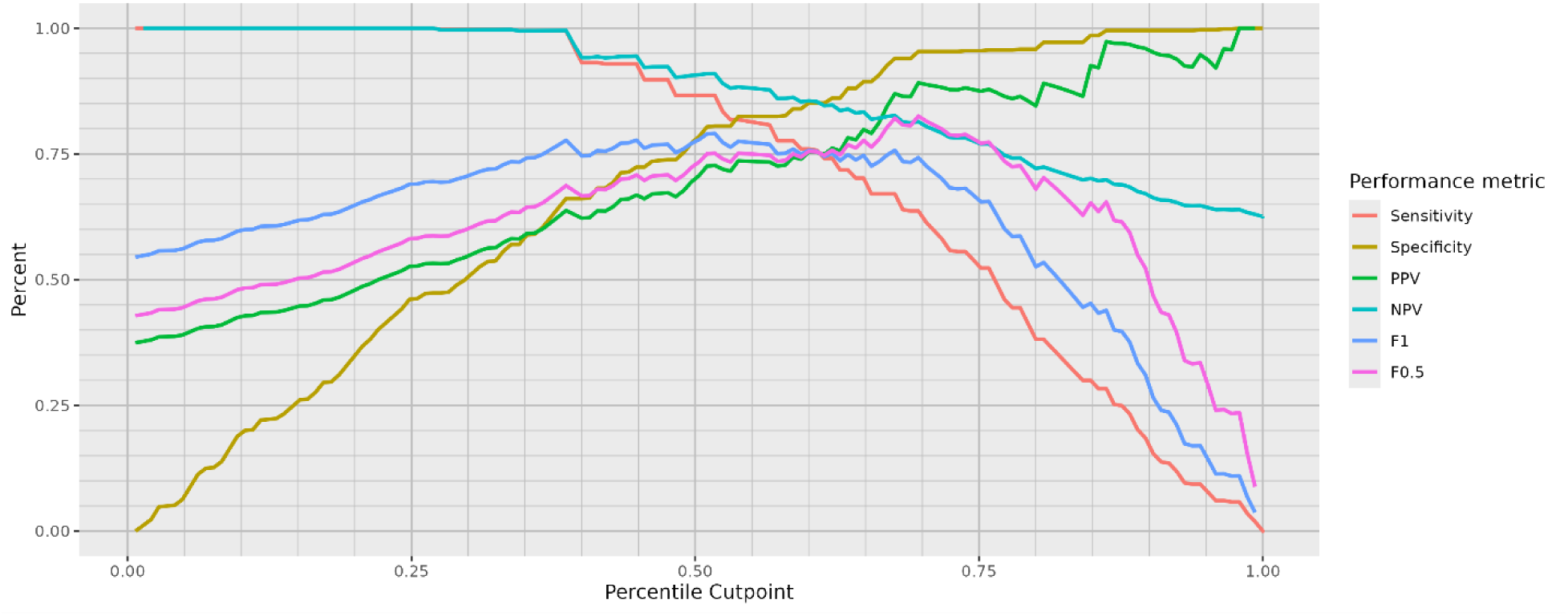
Model performance metrics at different cutpoints of predicted risk for the aggregate model trained at VUMC (using 1059 episodes without gold-standard labels) and evaluated on 145 episodes with gold-standard labels from KPWA. The horizontal axis is the quantile of the distribution of model-predicted probability.

**Figure 4:**
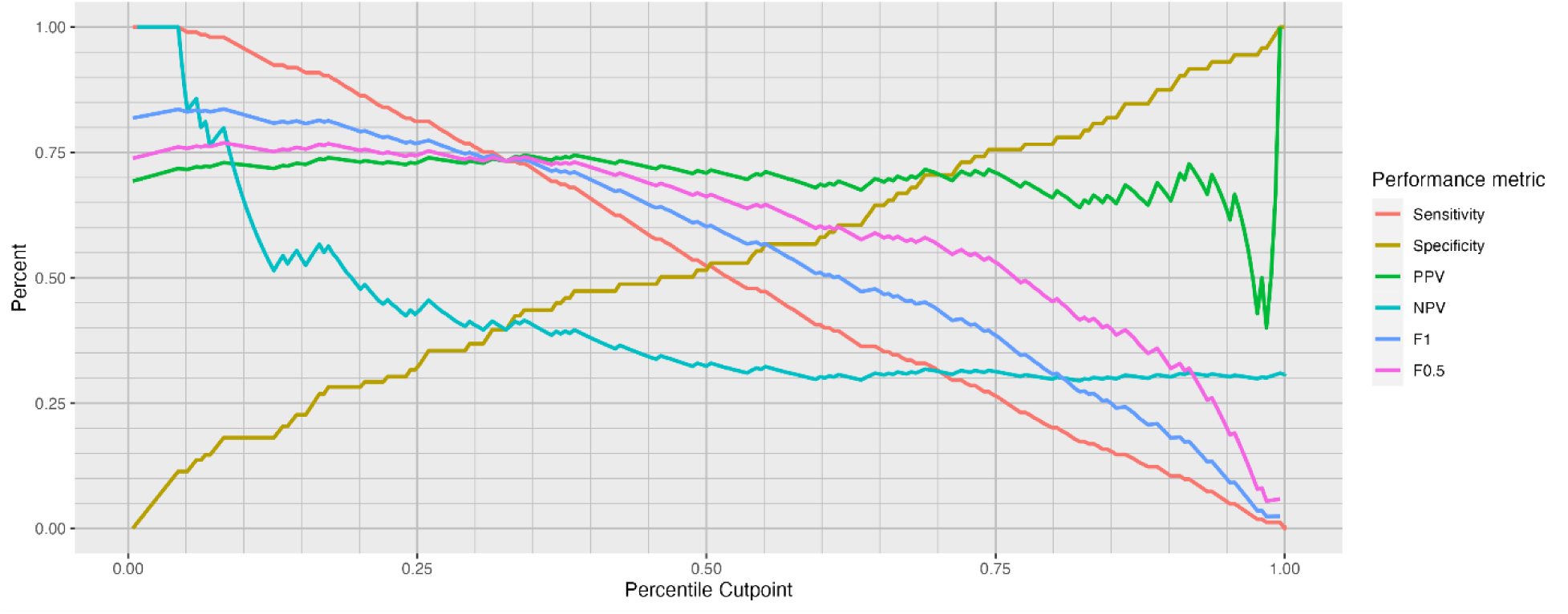
Model performance at different cutpoints of predicted risk for the aggregate model trained at KPWA (using 883 episodes without gold-standard labels) and evaluated on 254 episodes with gold-standard labels from VUMC. The horizontal axis is the quantile of the distribution of model-predicted probability.

### Prediction performance among vaccine-proximal and non-proximal episodes at KPWA

We observed slight differences in prediction performance between vaccine-proximal and non-proximal episodes at KPWA (Table 4). Prediction performance tended to be better among non-proximal episodes overall, with AUCs ranging from 0.687 to 0.938, compared to AUCs ranging from 0.621 to 0.773 among vaccine-proximal episodes. However, performance at specific thresholds could be higher among vaccine-proximal episodes, with sensitivity ranging from 0.93 to 1 at the cutoff that maximized F1 score compared to sensitivity for non-proximal episodes ranging from 0.79 to 0.95 at the cutoff that maximized F1.

**Table 4.** Performance of PheNorm models by presence of proximal vaccine at KPWA: Prediction performance of models trained at KPWA based on each silver label evaluated on the 145 episodes with gold-standard labels at KPWA, stratified by whether the index encounter was one of the 40 episodes proximal to a vaccine (defined as a vaccine two days prior to the encounter through the day of the encounter).

| Proximal to vaccine? | Silver label | AUC [95% CI] | Cutoff | Quantile | Sensitivity | Specificity | PPV | NPV | F<sub>1</sub> | F<sub>0.5</sub> |
| --- | --- | --- | --- | --- | --- | --- | --- | --- | --- | --- |
| No (n = 105) | Anaphylaxis diagnosis codes<sup>1</sup> | 0.687 [0.586, 0.787] | 0.4990 | 0.33 | 0.92 | 0.51 | 0.53 | 0.91 | 0.67 | 0.58 |
|  |  |  | 0.4987 | 0.30 | 0.95 | 0.47 | 0.52 | 0.94 | 0.67 | 0.57 |
|  | Anaphylaxis mentions | 0.896 [0.841, 0.952] | 0.6866 | 0.56 | 0.82 | 0.85 | 0.77 | 0.89 | 0.79 | 0.78 |
|  | Anaphylaxis CUIs | 0.878 [0.814, 0.943] | 0.8939 | 0.59 | 0.79 | 0.87 | 0.78 | 0.87 | 0.78 | 0.78 |
|  | Anaphylaxis or epinephrine mentions | 0.938 [0.895, 0.981] | 0.2187 | 0.49 | 0.93 | 0.79 | 0.73 | 0.95 | 0.82 | 0.76 |
|  | Aggregate | 0.909 [0.857, 0.960] | 0.6153 | 0.53 | 0.85 | 0.83 | 0.75 | 0.90 | 0.80 | 0.77 |
| Yes (n = 40) | Anaphylaxis diagnosis codes | 0.621 [0.443, 0.800] | 0.4956 | 0.23 | 0.93 | 0.28 | 0.44 | 0.88 | 0.60 | 0.49 |
|  | Anaphylaxis mentions | 0.765 [0.623, 0.908] | 0.0037 | 0.33 | 1.00 | 0.48 | 0.54 | 1.00 | 0.70 | 0.59 |
|  | Anaphylaxis CUIs | 0.752 [0.605, 0.899] | 0.0173 | 0.35 | 1.00 | 0.52 | 0.56 | 1.00 | 0.71 | 0.61 |
|  | Anaphylaxis or epinephrine mentions | 0.773 [0.631, 0.915] | 0.3427 | 0.55 | 0.80 | 0.72 | 0.63 | 0.86 | 0.71 | 0.66 |
|  | Aggregate | 0.768 [0.625, 0.911] | 0.1330 | 0.35 | 1.00 | 0.52 | 0.56 | 1.00 | 0.71 | 0.61 |

<sup>1</sup>Multiple rows are shown for this model because multiple cutoffs achieved F1 score within 0.0005 of each other.

## Discussion

Our results demonstrate that PheNorm can be applied successfully to develop a computable phenotyping algorithm to identify anaphylaxis in electronic healthcare data. AUCs ranged from 0.684—0.931 when evaluated on KPWA data and ranged from 0.521—0.673 when evaluated on VUMC data. Sensitivity at the maximum F1 score for the best performing models was greater than 90%, while PPV was moderate (near 70%). Sensitivity tended to be higher for more cutpoints in the KPWA evaluation data, while PPV tended to be higher for more cutpoints in the VUMC evaluation data. These results are better than expected, because anaphylaxis has high clinical and data complexity, often leading to poor algorithm prediction performance ^2,39^. Our best-performing models had better PPV and sensitivity tradeoffs than prior bespoke anaphylaxis models with costly, manually curated features.

Performance varied by silver label and evaluation dataset. The anaphylaxis diagnosis silver label model had consistent AUC (0.67—0.75) and high sensitivity (>0.87) regardless of model training site and evaluation site. NLP-based silver label models tended to perform much better at KPWA (AUC > 0.87) than at VUMC (AUC ≤0.60). Both models (those trained at VUMC and those trained at KPWA) had the same top-performing and worst-performing silver labels when evaluated on the same dataset. Future work should use the model (i.e., one of the four silver label-specific models or the aggregate model) that makes appropriate tradeoffs for the scientific context. The performance of the models we developed may be acceptable in certain scientific investigations, such as vaccine safety surveillance, where all suspected cases of anaphylaxis are traditionally manually abstracted and then reviewed by health care providers. For example, using the KPWA aggregate model and evaluated on KPWA data, we can achieve 94% sensitivity at a model-predicted probability quantile of 0.3, implying that using this threshold would identify most true cases.

The variation in performance between healthcare system sites is likely due to the population of eligible encounters at KPWA differing from those at VUMC in one key aspect. Because KPWA does not have inpatient or emergency facilities, care in these contexts is captured through claims alone. Thus all inpatient or ED encounters (path 1 and 3) at KPWA had to have an outpatient follow-up appointment (or engage in secure messaging with their care team) that generated note(s) with sufficient text. Approximately 40% of initial index encounters were excluded for insufficient text at KPWA compared to less than 1% at VUMC. The NLP output utilized by PheNorm at KPWA included concepts from only notes generated from outpatient visits or secure messaging; at VUMC, PheNorm utilized CUIs from all inpatient, outpatient, and ED notes. For inpatient encounters especially, anaphylaxis was rarely the focus of the encounter and thus may not have been documented frequently in the notes. Nearly 50% of encounters at VUMC involved >4 days with notes (Table 1), highlighting the complexity and possible heterogeneity of notes at VUMC. Information related to other aspects of the encounter may thus have obscured the anaphylaxis signal for the NLP-based silver labels compared to KPWA.

The KPWA models also performed differently for encounters near a vaccine compared to those not near a vaccine, tending to have worse AUC but higher sensitivity among vaccine-proximal encounters. This difference may be influenced by the fact that all encounters proximal to a vaccine were reviewed and thus were not available for model training. Vaccine status was not captured at VUMC.

Our work has several limitations. First, as mentioned above, the eligible encounters at KPWA and VUMC differ because KPWA does not have inpatient or emergency facilities, so these two systems represent different populations of potential anaphylaxis encounters. Second, we used data from only two healthcare systems; while geographically and demographically diverse, these systems may not be representative of other settings in the US. Finally, our approach requires access to text from clinical notes, which may not be available in all settings.

## Conclusion

PheNorm was used to successfully develop a computable phenotyping algorithm to identify anaphylaxis, an acute health condition that has both high data complexity and clinical complexity, using electronic healthcare data. The best-performing models had better PPV and sensitivity tradeoffs than prior bespoke anaphylaxis models using manually curated features. In this study, model performance varied widely depending on the silver label used and on the health care system used for evaluation. At VUMC, the anaphylaxis diagnosis code silver label model had the largest AUC, while NLP-based silver label models performed poorly. At KPWA, where all notes came from follow-up encounters, the diagnosis code silver label model had the smallest AUC, while the anaphylaxis or epinephrine mentions model had the largest AUC. Both models performed similarly on the same evaluation data, providing preliminary evidence that the models are transportable despite key differences in the populations giving rise to the data. Further research on using PheNorm for identification of anaphylaxis is necessary, especially comparing sites with varying access to hospital and emergency department data.

## Supporting information

Supplemental Section 2.2 MedDRA mapping

Supplemental Section 1.3 Medical record abstraction form instructions

Supplemental Section 1.3 Medical record abstraction form

## Acknowledgments

We thank the members of the Sentinel Innovation Center Workgroup and the Vaccine Safety Methods Working Group that provided helpful feedback during development of this work.

## Author contributions

BDW, JCS, DSC, and JCN conceptualized and led the study at KPWA (BDW, DSC, JCN) and VUMC (JCS). BDW, JCS, DSC, and JCN contributed to the design and planning of study methodology. Data acquisition and analysis were performed by JCS, DP, RW, JW, MM, DJC, OY, AR, SF, JC, EK, and BDW. Chart review was performed by JW, MM, and JC. BDW and JCS wrote the initial draft; all authors contributed revisions and approved the final version of the manuscript.

## Supplementary material

Supplementary material, including tables defining the anaphylaxis paths from Methods and figures with additional results, is available online.

## Data availability

The data underlying this article cannot be shared publicly due to institutional policies that protect the privacy of individuals whose data was used in the study.

