## Supplemental Section 2.2 MedDRA mapping for "Identifying anaphylaxis using weakly-supervised prediction models and natural language processing"

| LLT-CUI | LLT | PT-CUI | PT |
|---|---|---|---|
| C0000729 | Griping abdomen | C0000729 | Abdominal Pain |
| C0221512 | Stomach ache | | |
| C0232488 | Abdominal colic | | |
| C0232491 | Chronic abdominal pain | | |
| C0344304 | Abdominal pain generalized | | |
| C0423644 | Central abdominal pain | | |
| C0522061 | Abdominal pain localized | | |
| C0549273 | Abdominal pain aggravated | | |
| C0857092 | Duodenal ulcer-type symptoms | | |
| C0857334 | Colicky | | |
| C0920154 | Spleen pain | | |
| C1096624 | Periumbilical pain | | |
| C1609533 | Functional abdominal pain | | |
| C1662979 | Peritoneal pain | | |
| C1698089 | Postprandial pain | | |
| C3805159 | Mesogastric pain | | |
| C0340865 | Anaphylactoid shock | C0000729 | anaphylaxis |
| C0344159 | Anaphylactic reaction to venom | | |
| C0344183 | Exercise-induced anaphylaxis | | |
| C0685898 | Anaphylactic shock due to adverse food | | |
| C0857035 | Acute anaphylaxis | | |
| C1096621 | Anaphylactic reaction to chemical | | |
| C1536101 | Anaphylactic reaction to vaccine | | |
| C5441893 | Anaphylactic reaction to drug | | |
| C5442289 | Anaphylactic reaction to contrast agent | | |
| C5577808 | Biphasic anaphylactic reaction | | |
| C0549440 | Angioneurotic oedema aggravated | | |
| C0743747 | Face angioedema | | |
| C0857036 | Acute angio oedema | | |
| C0857323 | Angioedema of larynx | | |
| C0859055 | Oedema vascular | | |

| | | |
|---|---|---|
| C0863128 Angioedema aggravated<br>C0877772 Angioneurotic edema, not elsewhere cla<br>C0920175 Allergic angioedema<br>C1304200 Lip angioedema<br>C4324658 Eye angioedema<br>C4324659 Respiratory angioedema | **C0002994** | **Angioedema** |
| C0154455 Other anxiety states<br>C0233481 Worry<br>C0233485 Apprehension<br>C0235107 Anxiety-ridden<br>C0235111 Anxiety complex<br>C0262377 Situational anxiety<br>C0349231 Phobia, unspecified<br>C0542180 Reaction alarm<br>C0549258 Angor animi<br>C0549259 Anxiety aggravated<br>C0581386 Chronic anxiety<br>C0683278 Anguish<br>C0700031 Anxiety attack<br>C0700613 Anxiety state<br>C0853074 Anxiety NEC<br>C0853241 Exacerbation of anxiety<br>C0857102 Feeling of intense apprehension<br>C0857126 Impending doom<br>C0857480 Made anxiety worse<br>C0859031 Face anguish<br>C0860603 Anxiety symptoms<br>C2239195 Anxious mood<br>C2945761 Alarm (not alarm reaction) | **C0003467** | **Anxiety** |
| C0155877 Atopic asthma<br>C0155878 Extrinsic asthma without mention of stat<br>C0155880 Intrinsic asthma | | |

| | | |
|---|---|---|
| C0155881 Intrinsic asthma without mention of stat<br>C0155886 Asthma, unspecified type, without ment<br>C0340067 Drug-induced asthma<br>C0340076 Eosinophilic asthma<br>C0347950 Asthmatic attack<br>C0349790 Exacerbation of asthma<br>C0549336 Asthma aggravated<br>C0858626 Asthmatic attack induced<br>C0859987 Asthmatoid bronchitis<br>C0877430 Asthma chronic<br>C0877446 Asthma-like condition<br>C0948683 Asthmatic attack atopic<br>C1273489 Nocturnal asthma<br>C1319018 Wheezy bronchitis<br>C1735623 Cold induced asthma<br>C2363774 Neutrophilic asthma<br>C2363819 Paucigranulocytic asthma<br>C2747883 Infection induced asthma<br>C2919352 Seasonal asthma<br>C5394677 Thunderstorm asthma | **C0004096** | **Asthma** |
| C0015263 Exercise-induced bronchospasm<br>C0079043 Bronchoconstriction<br>C0235906 Bronchospasm aggravated<br>C1609536 Allergic bronchospasm | **C0006266** | **Bronchospasm** |
| C0029537 Other chest pain<br>C0151826 Substernal chest pain<br>C0232286 Chest pain precordial<br>C0232288 Chest pain exertional<br>C0235710 Chest discomfort<br>C0235715 Chest aching of<br>C0235716 Chest burning pain of<br>C0241243 Sternal pain | | |

| | | | |
|---|---|---|---|
| C0302355 | Chest pain NEC | C0008031 | Chest Pain |
| C0522051 | Acute chest pain | | |
| C0541828 | Chest pain-L arm | | |
| C0740396 | Chest burning | | |
| C0856134 | Chest pain aggravated | | |
| C0856578 | Parasternal pain (excluding organic) | | |
| C0857030 | Ache across chest | | |
| C0857367 | Generalized chest pain | | |
| C1112757 | Chest pain with radiation to left arm | | |
| C1740831 | Chronic chest pain | | |
| C3203733 | Precordial catch syndrome | | |
| C0152124 | Reactive confusion | C0009676 | Confusion |
| C0154333 | Subacute confusional state | | |
| C0338642 | Toxic confusional state | | |
| C0813178 | Bewilderment | | |
| C0856071 | Confusion aggravated | | |
| C0856480 | Confusional state (organic, acute or suba | | |
| C0857042 | Acute onset of confusion | | |
| C0857386 | Nocturnal confusion | | |
| C0858851 | Confusion reversible | | |
| C1285577 | Acute confusion state | | |
| C0010201 | Chronic cough | C0010200 | Coughing |
| C0231911 | Paroxysmal cough | | |
| C0231912 | Nocturnal cough | | |
| C0264345 | Smoker's cough | | |
| C0423730 | Painful cough | | |
| C0455777 | Cough ineffective | | |
| C0562483 | Persistent cough | | |
| C0574067 | Cough increased | | |
| C0742857 | Acute cough | | |
| C0850149 | Dry cough | | |
| C0856112 | Cough aggravated | | |

| | | |
|---|---|---|
| C0857172 Persistent dry cough<br>C0857173 Persistent non-productive cough<br>C0857513 Cough resembling asthma<br>C0859870 Irritative cough<br>C1096640 Coughing after drug inhalation<br>C1142295 Drug-induced cough<br>C2202983 Cough weak<br>C4552485 Unexplained chronic cough<br>C4552486 Refractory chronic cough | | |
| C0156173 Functional diarrhea<br>C0239182 Watery diarrhoea<br>C0267556 Osmotic diarrhea<br>C0267557 Secretory diarrhea<br>C0401151 Chronic diarrhea<br>C0578159 Antibiotic-associated diarrhea<br>C0740441 Acute diarrhea<br>C0853947 Diarrhoea aggravated<br>C0856217 Functional diarrhoea (due to spastic colo<br>C0857098 Explosive diarrhoea<br>C0857138 Frank diarrhoea<br>C0857184 Urgent diarrhoea<br>C0859971 Idiopathic diarrhoea<br>C0859975 Nocturnal diarrhoea<br>C0860247 Iatrogenic diarrhoea<br>C0860527 Mushy stool<br>C1868939 Mushy diarrhea<br>C2129214 Loose bowels<br>C2747921 Malodorous diarrhea<br>C3671554 Mucous diarrhea | **C0011991** | **Diarrhea** |
| C0039070 Syncope<br>C0220870 Lightheadedness<br>C0476206 Dizziness and giddiness | | |

| | | |
|---|---|---|
| C0476207 Giddiness<br>C0549239 Dizziness aggravated<br>C0581879 Felt faint<br>C0595963 Wooziness<br>C0852858 Vertigo (excluding dizziness)<br>C0857087 Dizzy spells<br>C0857226 Swaying feeling<br>C0857314 Felt giddy<br>C5442487 Idiopathic dizziness | **C0012833** | **Dizziness** |
| C0030824 Penicillin allergy<br>C0038757 Sulfonamide allergy<br>C0571622 Insulin allergy<br>C0741103 Allergy to antibiotic<br>C0850093 Allergic reaction to antibiotics<br>C0857594 Specific allergy (drug)<br>C3267191 Allergic reaction to analgesics<br>C4708098 Allergy to vitamin B12<br>C4727850 Allergy to topical drugs<br>C5400282 Allergy to ayurvedic drugs<br>C5400285 Allergy to allopathic drugs<br>C5400286 Allergy to homeopathic drugs<br>C5442512 Allergic reaction to chemotherapy<br>C5553031 Allergy to NSAIDs | **C0013182** | **Drug Allergy** |
| C0029601 Other dyspnea and respiratory abnorma<br>C0159053 Dyspnea and respiratory abnormalities<br>C0231848 Air hunger<br>C0425449 Gasping<br>C0553668 Labored breathing<br>C0574066 Increased shortness of breath<br>C0743323 Acute dyspnea<br>C0853326 Dyspnoea exacerbated<br>C0857142 Marked inactivity of chest wall on inspira | **C0013404** | **Dyspnea** |

| | | |
|---|---|---|
| C0859927 Increased work of breathing<br>C0860550 Respiratory tract closed sensation of | | |
| C0234940 Inflammatory oedema reaction<br>C0238094 Idiopathic edema<br>C0333241 Chronic edema<br>C0333243 Pitting edema<br>C0333244 Transient edema<br>C0856110 Oedema aggravated<br>C0857441 Oedematous weight gain<br>C0858531 Oedema-like<br>C2938876 Weeping edema | **C0013604** | **Edema** |
| C0014238 Parasitic endophthalmitis NOS<br>C0029610 Other endophthalmitis<br>C0154773 Acute endophthalmitis<br>C0154774 Chronic endophthalmitis<br>C0259800 Purulent endophthalmitis<br>C0859152 Other specified endophthalmitis<br>C1532322 Infectious endophthalmitis | **C0014236** | **Endophthalmitis** |
| C0159038 Pallor and flushing<br>C0856089 Flushing aggravated<br>C0857115 Gross flushing<br>C0857370 Generalized flushing<br>C0860135 Flushed chest | **C0016382** | **Flushing** |
| C0559469 Egg allergy<br>C0559470 Peanut allergy<br>C0577620 Allergy to nuts<br>C0577625 Shellfish allergy<br>C0685900 Seafood allergy<br>C0685901 Fruit allergy<br>C0856904 Fish allergy<br>C0856905 Potato allergy<br>C0857593 Ovalbumin allergy | | |

| | | | |
|---|---|---|---|
| C0859949 | Meat allergy | C0016470 | Food Allergy |
| C1096643 | Allergy to grains | | |
| C1096735 | Allergy to legumes | | |
| C1112677 | Protein allergy | | |
| C1630645 | Alpha-gal allergy | | |
| C1690580 | Allergic reaction to food | | |
| C2363729 | Spice allergy | | |
| C2938930 | Food allergen sensitisation | | |
| C4075590 | Soy allergy | | |
| C4082933 | Allergy to animal | | |
| C4552514 | Vegetable allergy | | |
| C4761189 | Allergy to edible fungus | | |
| C5400281 | Oil allergy | | |
| C5577936 | Sesame allergy | | |
| C0857375 | Hypoxic arrest | C0018790 | Cardiac Arrest |
| C0858523 | Cardiac arrest transient | | |
| C0858981 | Congestive cardioplegia | | |
| C4285916 | Ventricular standstill | | |
| C0235699 | Red neck syndrome | C0020517 | Hypersensitivity |
| C0235893 | Allergy aggravated | | |
| C0282504 | Environmental allergy | | |
| C0375321 | Upper respiratory tract hypersensitivity r | | |
| C0413234 | Acute allergic reaction | | |
| C0869403 | Allergy, unspecified, not elsewhere class | | |
| C1527304 | Allergic reaction | | |
| C1736167 | Systemic allergic reaction | | |
| C0155800 | Chronic hypotension | | |
| C0375314 | Iatrogenic hypotension | | |
| C0476454 | Blood pressure reading low | | |
| C0520541 | Hypotensive episode | | |
| C0740481 | Hypotension asymptomatic | | |
| C0745176 | Acute hypotension | | |

| | | |
|---|---|---|
| C0856090 Hypotension aggravated<br>C0857353 Hypotensive<br>C0858763 Hypotension paroxysm<br>C0858766 Blood pressure dropped transient<br>C0861177 Mean blood pressure decreased<br>C0863113 Hypotension symptomatic<br>C0948686 Preshock<br>C1699115 Transient systolic hypotension | **C0020649** | **Hypotension** |
| C0021381 Exudative inflammation<br>C0234939 Inflammatory swelling<br>C0522570 Inflammation localized | **C0021368** | **Inflammation** |
| C0021925 Intubation<br>C0857231 To facilitate intubation<br>C0860359 Reintubate<br>C1868878 Endotracheal reintubation | **C0021932** | **Endotracheal intubation** |
| C0201682 Chemistry NOS | **C0022885** | **Laboratory Procedures** |
| C0235552 Subglottic edema<br>C0264315 Glottic edema<br>C0392319 Vocal cord edema<br>C0472519 Reinke's edema<br>C1142210 Aryepiglottis oedema<br>C1735912 Acute laryngeal edema<br>C5554320 Pharyngolaryngeal edema | **C0023052** | **Laryngeal Edema** |
| C1112473 Mast cell aggregation | **C0024899** | **Mastocytosis** |
| C0375548 Nausea alone<br>C0549280 Nausea aggravated<br>C0746783 Postprandial nausea<br>C0857071 Churning of stomach<br>C0857218 Sickness/nausea<br>C1112312 Nausea post chemotherapy<br>C3495909 Nausea post radiotherapy | **C0027497** | **Nausea** |

| | | |
|---|---|---|
| C3854611 Anticipatory nausea | | |
| C0085668 Secondary carcinoma<br>C0153675 Secondary malignant neoplasm of respir<br>C0153684 Secondary malignant neoplasm of other<br>C0856617 Secondary carcinoma (known primary)<br>C0859898 Secondary malignant neoplasm of other<br>C5554011 Synchronous metastases<br>C5554406 Metachronous metastasis | **C0027627** | **Neoplasm Metastasis** |
| C0030200 Intractable pain<br>C0150055 Chronic pain<br>C0184567 Acute pain<br>C0234230 Pain burning<br>C0234238 Ache<br>C0234250 Referred pain<br>C0234252 Mechanical pain<br>C0234253 Pain at rest<br>C0234254 Radiating pain<br>C0235048 Smarting<br>C0278144 Pain dull<br>C0278146 Shooting pain<br>C0278148 Throbbing pain<br>C0281856 General body pain<br>C0423603 Pricking pain<br>C0520962 Pain localized<br>C0520963 Diffuse pain<br>C0542135 pain left side<br>C0677500 Stinging<br>C0853946 Pain exacerbated<br>C0858802 Burn lesion pain transient aggravation of<br>C0858803 Pain irritated<br>C0858805 Prick pain feeling<br>C0948843 Irradiating pain | **C0030193** | **Pain** |

| | |
|---|---|
| C0948844 Lancinating pain<br>C1317590 Trigger point pain<br>C2363726 Pain upon movement<br>C3805255 Residual pain<br>C4087510 Kinesalgia<br>C4523910 Pain post chemotherapy<br>C4524193 Pantalgia | |
| C0039231 Tachycardia<br>C0235240 Heart pounding<br>C0549267 Palpitations aggravated<br>C4727920 Exercise induced palpitation | **C0030252** **Palpitations** |
| C0157724 Pruritus and related conditions<br>C0157725 Other specified pruritic conditions<br>C0239653 Itchy feet<br>C0240941 Itchy scalp<br>C0475858 Generalized pruritus<br>C0856060 Pruritus aggravated<br>C0857130 Itchy legs<br>C0858706 Extremities itchy sensation of<br>C0858708 Itch burning<br>C0858709 Pruritus of both hands<br>C0919793 Pruritus breast<br>C0948847 Hairy skin itching<br>C1142396 Localized itching<br>C3670839 Pruritus facial<br>C4523818 Itchy upper limbs<br>C5208084 Periorbital pruritus<br>C5243975 Localised pruritus<br>C5577868 Chronic pruritus of unknown origin | **C0033774** **Pruritus** |
| C0007203 Cardiopulmonary resuscitation | **C0035273** **Resuscitation procedure** |
| C0029737 Other shock without mention of trauma | |

| | | |
|---|---|---|
| C0157450 Shock during or following labor and deliv | C0036974 | Shock |
| C0157451 Shock during or following labor and deliv | | |
| C0157452 Shock during or following labor and deliv | | |
| C0157453 Shock during or following labor and deliv | | |
| C0157454 Shock during or following labor and deliv | | |
| C0157455 Shock during or following labor and deliv | | |
| C0159051 Shock without mention of trauma | | |
| C0221477 Vasomotor collapse | | |
| C0344329 Collapse | | |
| C0349412 Refractory shock | | |
| C0495170 Shock following abortion and ectopic and | | |
| C0853340 Peripheral shutdown | | |
| C0856621 Shock (excluding traumatic and specific c | | |
| C0856661 Clinically shocked | | |
| C0857075 Collapsed semi conscious & shocked | | |
| C0859211 Shock during or following labor and deliv | | |
| C0859274 Postoperative shock, not elsewhere class | | |
| C0749874 Upper respiratory symptom | **C0037090** | **Respiratory symptom** |
| C0021881 Intracutaneous test | C0037296 | Hypersensitivity skin testing |
| C0030646 Skin patch test | | |
| C0430561 Prick test | | |
| C5577824 PEG skin test | | |
| C5577825 Polyethylene glycol skin test | | |
| C0677600 Stridor inspiratory | **C0038450** | **Stridor** |
| C0159045 Swelling, mass, or lump in head and neck | C0038999 | Swelling |
| C0235434 Swelling non-inflammatory | | |
| C0235435 Tissue puffing | | |
| C0423601 Feeling swollen | | |
| C0476228 Localized superficial swelling, mass, or lu | | |
| C0476313 Groin swelling | | |
| C0541781 Arms swollen red hot | | |

| | | |
|---|---|---|
| C0578454 Neck swelling<br>C0749356 Swelling femoral<br>C0853619 Local swelling<br>C0858956 Swelling of dorsum manus | | |
| C0042420 Vasovagal syncope<br>C0234434 Syncope hypotensive<br>C0234435 Cough syncope<br>C0234437 Syncope postural<br>C0235242 Syncope exertional<br>C0340850 Neurally mediated syncope<br>C0340855 Syncope micturition<br>C0340856 Defecation syncope<br>C0749201 Orthostatic syncope<br>C0751535 Cardiac syncope<br>C0751536 Syncope convulsive<br>C0856092 Syncope aggravated<br>C0877090 Orthostatic collapse<br>C2721582 Swallow syncope<br>C3854672 Arrhythmic syncope<br>C4049594 Cardiovascular syncope | **C0039070** | **Syncope** |
| C0549268 Tachycardia aggravated<br>C0749249 Wide complex tachycardia<br>C0856629 Tachycardia (excluding paroxysmal)<br>C0858524 Tachycardia nervous<br>C0858871 Heart sound accelerated<br>C1328539 Reflex tachycardia | **C0039231** | **Tachycardia** |
| C0312422 Blacked out<br>C0853202 Loss of consciousness NEC<br>C4087400 Transient loss of consciousness | **C0041657** | **Unconscious State** |
| C0413402 Adverse reaction to antibiotics | **C0041755** | **Adverse reaction to drug** |
| C0029839 Other specified urticaria | | |

| | | |
|---|---|---|
| C0149526 Allergic urticaria<br>C0221232 Welts<br>C0234935 Acute urticaria<br>C0263347 Urticaria drug-induced<br>C0750016 Generalized urticaria<br>C0853333 Urticaria aggravated<br>C0853691 Histamine-like rash<br>C0854430 Urticaria subcutaneous<br>C0856779 Acute allergic urticaria<br>C0856780 Pharmacological urticaria<br>C0857390 Generalized urticarial rash<br>C0858686 Rash urticaria-like<br>C0858687 Urticarial symptom<br>C0859045 Erythema urticarial<br>C1261981 Urticaria localised<br>C1276118 Urticaria recurrent<br>C1536459 Nettle rash<br>C4728195 Atopic urticaria | **C0042109** | **Urticaria** |
| C0027498 Nausea and vomiting<br>C0151791 Nausea vomiting and diarrhea<br>C0152165 Persistent vomiting<br>C0232599 Bilious vomiting<br>C0235250 Hyperemesis<br>C0267172 Habit vomiting<br>C0728950 Vomiting alone<br>C0856095 Vomiting aggravated<br>C0859028 Vomiting reflex<br>C0948239 Vomiting of medication<br>C0948394 Vomiting post chemotherapy<br>C0949067 Tablet in vomitus<br>C1112670 Vomiting post radiotherapy<br>C1142579 Blennemesis | **C0042963** | **Vomiting** |

| | | |
|---|---|---|
| C1168325 Postprandial emesis | | |
| C0231874 Wheezing inspiratory<br>C0231875 Wheezing expiratory<br>C0392681 Asthmatic wheezing<br>C0857522 Increased wheeziness<br>C0877638 Wheezing aggravated<br>C1168179 Sibilus<br>C1735643 Chronic wheezing | **C0043144** | **Wheezing** |
| C0266815 Cow's milk allergy<br>C3889086 Milk protein allergy<br>C4758639 Cow's milk protein allergy | **C0079840** | **Milk Allergy** |
| C0814177 Noninvasive procedure<br>C0854626 Ulcer management<br>C2363849 Non-surgical treatment<br>C4048276 Invasive procedure | **C0087111** | **Therapeutic procedure** |
| C0238977 Cheek swelling<br>C0856911 Swelling lips & face<br>C0857229 Swelling of face & neck<br>C0858703 Swollen of face feeling<br>C1112766 Forehead swelling<br>C4049344 Eyebrow swelling<br>C4049484 Swelling of jaw angle<br>C4280766 Jaw swelling<br>C5208051 Temple swelling | **C0151602** | **Facial swelling** |
| C0856668 Face & tongue oedema | **C0151610** | **Edema of the tongue** |
| C0863180 Respiratory arrest (excluding neonatal) | **C0162297** | **Respiratory arrest** |
| C0033144 Primary prevention<br>C0445202 Prophylactic<br>C0679699 Secondary prevention<br>C0679700 Tertiary prevention | **C0199176** | **Prophylactic treatment** |
| C0554804 Assisted ventilation | | |

| | | |
|---|---|---|
| C1868981 Invasive mechanical ventilation<br>C1868982 Noninvasive mechanical ventilation<br>C4760774 Transnasal humidified rapid-insufflation | **C0199470** | **Mechanical ventilation** |
| C0220854 Hyperpnea<br>C0858639 Respiration stimulated | **C0231835** | **Tachypnea** |
| C1504512 Medication aspiration | **C0232070** | **Foreign body aspiration** |
| C0232285 Chest pressure sensation of<br>C0232292 Chest tightness<br>C0235711 Chest distressed feeling of<br>C0235712 Chest fullness of<br>C0438716 Chest pressure<br>C0742339 Chest heaviness<br>C0856135 Chest tightness aggravated<br>C0857399 Retrosternal burning<br>C0857400 Retrosternal discomfort<br>C0858787 Chest abnormal feeling of<br>C0858788 Chest anxiety feeling of<br>C0858789 Chest hot feeling of<br>C0858790 Chest strangled feeling of<br>C1142568 Chest pressure exertional<br>C5400139 Chest wall discomfort | **C0235710** | **Chest discomfort** |
| C0858939 Pharynx closed sensation of | **C0236071** | **Constriction in throat** |
| C0236066 Lips swelling non-specific | **C0240211** | **Lip swelling** |
| C0155910 Pulmonary congestion and hypostasis<br>C0582411 Pulmonary venous congestion<br>C0858975 Pulmonary configuration increased<br>C1262246 Pulmonary stasis<br>C1737267 Acute pulmonary congestion | **C0242073** | **Pulmonary congestion** |
| C0700292 Hypoxemia<br>C3203358 Hypoventilation<br>C5400503 Transient nocturnal oxygen desaturation | **C0242184** | **Hypoxia** |

| | | | |
|---|---|---|---|
| C5554231<br>C5554263<br>C5554264 | Refractory desaturation<br>Happy hypoxia<br>Silent hypoxia | **C0242184** | **Hypoxia** |
| C1142195 | Delayed anaphylactoid reaction | **C0340865** | **Anaphylactoid Reaction** |
| C0859943<br>C0859957<br>C1504524 | Hereditary allergy<br>Spontaneous allergy<br>Allergic shiner | **C0392707** | **Atopy** |
| C0236003 | Mucosal swelling | **C0521481** | **Mucous membrane edema** |
| C0856623<br>C0856910<br>C1096623<br>C4552534 | Orofacial oedema<br>Oedema lips & face<br>Maxillofacial oedema<br>Forehead edema | **C0542571** | **Facial edema** |
| C0549433<br>C1735594<br>C5577945 | Surgical intervention<br>Stereotactic surgery<br>Stab incision | **C0543467** | **Operative Surgical Procedures** |
| C0750310<br>C4523878 | Intravascular depletion<br>Extracellular fluid volume decreased | **C0546884** | **Hypovolemia** |
| C0001889<br>C0085628<br>C0151559<br>C0234439<br>C0235070<br>C0235071<br>C0424532<br>C0542207<br>C0683369<br>C0877450<br>C0877609<br>C1142529 | Akinetic mutism<br>Stupor<br>Central nervous system depression<br>Semi-coma<br>Unconscious partial<br>Arousal difficult<br>Semi-conscious<br>Sensorium decreased<br>Consciousness clouding<br>Obnubilation<br>Alertness decreased<br>Precoma | **C0549249** | **Depressed Level of Consciousness** |
| C0151885<br>C4761010 | Reaction aggravation<br>Adverse reaction to product | **C0559546** | **Adverse reactions** |

| | | | |
|---|---|---|---|
| C0235217 Skin vasodilatation<br>C0424829 Distended blood vessels<br>C0553725 Skin vasodilating<br>C0856757 Dilated veins<br>C1262231 Capillarectasia<br>C1328540 Peripheral vasodilatation<br>C1959622 Vein distended | | **C0595862** | **Vasodilation disorder** |
| C0241381 Throat burning sensation of<br>C0542075 Local throat irritation<br>C0562057 Burning in throat<br>C0858938 Pharynx burning sensation of<br>C0858940 Pharynx irritated sensation of<br>C0858941 Pharynx itchy sensation of<br>C0863112 Pharyngo-oral irritation<br>C4087337 Raw throat | | **C0700184** | **Throat irritation** |
| C0232071 Food aspiration<br>C0877412 Aspiration of gastrointestinal contents in<br>C2585629 Aspiration into trachea<br>C3665956 Vomit aspiration<br>C4324315 Silent aspiration<br>C4761191 Aspiration into bronchus | | **C0700198** | **Pulmonary aspiration** |
| C0159065 Other symptoms involving abdomen and<br>C0859796 Other specified symptoms involving abdo | | **C0740651** | **Abdominal symptom** |
| C4324301 Vocal cord spasm | | **C0859897** | **Vocal cord dysfunction** |
| C0375668 Late effect of adverse effect of drug, med<br>C0851307 Certain adverse effects not elsewhere cla<br>C0877761 Other specified adverse effect, not elsew | | **C0877248** | **Adverse event** |
| C0241955 Jellyfish sting | | **C1096052** | **Venomous sting** |
| C0034076 Pulmonary insufficiency following traum<br>C0035229 Respiratory insufficiency<br>C0340194 Respiratory failure type 1 | | | |

| | | | |
|---|---|---|---|
| C0398353 | Hypercapnic respiratory failure | C1145670 | Respiratory Failure |
| C0852895 | Respiratory failures (excluding neonatal) | | |
| C0877746 | Other pulmonary insufficiency, not elsew | | |
| C0948755 | Pulmonary failure | | |
| C2363912 | Respiratory failure aggravated | | |
| C3277226 | Restrictive respiratory insufficiency | | |
| C3805211 | Hypoxic respiratory failure | | |
| C4324356 | End stage lung disease | | |
| C0542098 | Nasal mucus increased | C1260880 | Rhinorrhea |
| C0558361 | Sniffles | | |
| C0858633 | Nasal discharge watery excessive | | |
| C1262304 | Gustatory rhinorrhoea | | |
| C0425481 | Respiratory sighs | C1260922 | Abnormal breathing |
| C0152229 | Unattended death | C1306577 | Death (finding) |
| C0277589 | Death unexplained | | |
| C0277608 | Hospital death | | |
| C0421619 | Dead on arrival | | |
| C0476463 | Found dead | | |
| C0857796 | Died in sleep | | |
| C1869004 | Death from natural causes | | |
| C3889051 | Death unascertained | | |
| C5400128 | Death by homicide | | |
| C0019825 | Hoarse voice | C1527344 | Dysphonia |
| C0029873 | Other voice disturbance | | |
| C0264614 | Hypernasality | | |
| C0264618 | Hyponasality | | |
| C0521007 | Hypophonia | | |
| C0751513 | Rhinolalia | | |
| C0854299 | Disturbance in loudness | | |
| C0857508 | Distorted voice | | |
| C0858567 | Phonation difficulty | | |

| | |
|---|---|
| C0858568 Voice lowered<br>C0860616 Resonance disorder<br>C0860618 Vocal tone disorder<br>C0860619 Vocal volume disorder<br>C1527340 Voice alteration | |
| C0021489 Intradermal injection<br>C0021490 Intralesional injection<br>C2957535 Intramuscular injection | **C1533685 Injection procedure** |
| C0856716 Asthma aspirin-sensitive<br>C1096641 Widal syndrome<br>C1328364 Analgesic asthma syndrome | **C3853540 Aspirin exacerbated respiratory disease** |
| C0375703 Anaphylactic shock due to unspecified fc<br>C0375707 Anaphylactic shock due to tree nuts and<br>C0391985 Anaphylactic shock, not elsewhere classi<br>C0858807 Drug shock<br>C0858808 Penicillin shock<br>C0859855 Anaphylactic shock due to peanuts<br>C0859856 Anaphylactic shock due to crustaceans<br>C0859857 Anaphylactic shock due to fruits and veg<br>C0859858 Anaphylactic shock due to fish<br>C0859859 Anaphylactic shock due to food additives<br>C0859860 Anaphylactic shock due to milk products<br>C0859861 Anaphylactic shock due to eggs<br>C3161345 Anaphylactic shock due to other specifie | **C4316895 Anaphylactic shock** |
