## Supplementary Material for "Identifying anaphylaxis using weakly-supervised prediction models and natural language processing"

### S1: Supplemental Methods

#### S1.1: Anaphylaxis encounter path eligibility criteria

Table S1: ICD-10 diagnosis codes for anaphylaxis.

| Code | Code description |
|---|---|
| T78.2XXA | Anaphylactic shock, unspecified, initial encounter |
| T88.6XXA | Anaphylactic reaction due to adverse effect of correct drug or medicament properly administered, initial encounter |
| T78.00XA | Anaphylactic reaction due to unspecified food |
| T78.01XA | Anaphylactic reaction due to peanuts |
| T78.02XA | Anaphylactic reaction due to shellfish (crustaceans) |
| T78.03XA | Anaphylactic reaction due to other fish |
| T78.04XA | Anaphylactic reaction due to fruits and vegetables |
| T78.05XA | Anaphylactic reaction due to tree nuts and seeds |
| T78.06XA | Anaphylactic reaction due to food additives |
| T78.07XA | Anaphylactic reaction due to milk and dairy products |

| | |
|---|---|
| T78.08XA | Anaphylactic reaction due to eggs |
| T78.09XA | Anaphylactic reaction due to other food products |
| T80.51XA | Anaphylactic reaction due to administration of blood/products, initial encounter |
| T80.52XA | Anaphylactic reaction due to vaccination, initial encounter |
| T80.59XA | Anaphylactic reaction due to other serum, initial encounter |
| T80.59XA | Anaphylactic reaction due to other serum, initial encounter |

Table S2: Diagnosis and procedure codes for symptoms, procedures, or treatments that may occur on the same calendar day as an anaphylaxis diagnosis.

| Group | Code | Code type | Code type and description |
|---|---|---|---|
| I | J9801 | ICD-10 | Bronchiospasm |
| | R061 | ICD-10 | Stridor |
| | J1200 | HCPCS | Injection of diphenhydramine |
| II | I959 | ICD-10 | Hypotension |
| | J0170 | HCPCS | Injection of epinephrine |
| | J0171 | HCPCS | Injection of epinephrine |
| | 92950 | CPT | CPR procedure |
| | 5A12012 | ICD-10 | CPR procedure |
| | 5A1221Z | ICD-10 | CPT procedure |

Table S3: ICD-10 diagnosis codes for allergy unspecified, other unspecified adverse effects of drugs, medicinal and biological substances in therapeutic use.

| Code | Code description |
|---|---|
| T50905A | Adverse effect of unspecified drugs, medicaments and biological substances, initial encounter |
| T410X5A | Adverse effect of inhaled anesthetics, initial encounter |
| T411X5A | Adverse effect of intravenous anesthetics, initial encounter |
| T41205A | Adverse effect of unspecified general anesthetics, initial encounter |

| | |
|---|---|
| T41295A | Adverse effect of other general anesthetics, initial encounter |
| T413X5A | Adverse effect of local anesthetics, initial encounter |
| T4145XA | Adverse effect of unspecified anesthetic, initial encounter |
| T8859XA | Other complications of anesthesia, initial encounter |
| T383X5A | Adverse effect of insulin and oral hypoglycemic [antidiabetic] drugs, initial encounter |
| T50995A | Adverse effect of other drugs, medicaments and biological substances, initial encounter |
| See Table S4 | The set of 164 distinct ICD-10 codes that map to ICD-9 code 995.29 (“Unspecified adverse effect of other drug, medicinal or biological substance”). |
| T78.40XA | Allergy, unspecified, initial encounter |
| T78.49XA | Other allergy, initial encounter |

Table S4: The set of 164 distinct ICD-10 codes that map to ICD-9 code 995.29 (“Unspecified adverse effect of other drug, medicinal or biological substance”).

| Code | Description |
|---|---|
| T360X5A | Adverse effect of penicillins, initial encounter |
| T361X5A | Adverse effect of cephalosporins and other beta-lactam antibiotics, initial encounter |
| T362X5A | Adverse effect of chloramphenicol group, initial encounter |
| T363X5A | Adverse effect of macrolides, initial encounter |
| T364X5A | Adverse effect of tetracyclines, initial encounter |
| T365X5A | Adverse effect of aminoglycosides, initial encounter |
| T366X5A | Adverse effect of rifampicins, initial encounter |
| T367X5A | Adverse effect of antifungal antibiotics, systemically used, initial encounter |
| T368X5A | Adverse effect of other systemic antibiotics, initial encounter |
| T3695XA | Adverse effect of unspecified systemic antibiotic, initial encounter |
| T370X5A | Adverse effect of sulfonamides, initial encounter |
| T371X5A | Adverse effect of antimycobacterial drugs, initial encounter |
| T372X5A | Adverse effect of antimalarials and drugs acting on other blood protozoa, initial encounter |
| T373X5A | Adverse effect of other antiprotozoal drugs, initial encounter |
| T374X5A | Adverse effect of anthelminthics, initial encounter |
| T375X5A | Adverse effect of antiviral drugs, initial encounter |
| T378X5A | Adverse effect of other specified systemic anti-infectives and antiparasitics, initial encounter |
| T3795XA | Adverse effect of unspecified systemic anti-infective and antiparasitic, initial encounter |
| T380X5A | Adverse effect of glucocorticoids and synthetic analogues, initial encounter |
| T381X5A | Adverse effect of thyroid hormones and substitutes, initial encounter |
| T382X5A | Adverse effect of antithyroid drugs, initial encounter |
| T384X5A | Adverse effect of oral contraceptives, initial encounter |
| T385X5A | Adverse effect of other estrogens and progestogens, initial encounter |
| T386X5A | Adverse effect of antigonadotrophins, antiestrogens, antiandrogens, NEC, initial encounter |
| T387X5A | Adverse effect of androgens and anabolic congeners, initial encounter |
| T38805A | Adverse effect of unspecified hormones and synthetic substitutes, initial encounter |
| T38815A | Adverse effect of anterior pituitary [adenohypophyseal] hormones, initial encounter |
| T38895A | Adverse effect of other hormones and synthetic substitutes, initial encounter |
| T38905A | Adverse effect of unspecified hormone antagonists, initial encounter |
| T38995A | Adverse effect of other hormone antagonists, initial encounter |
| T39015A | Adverse effect of aspirin, initial encounter |
| T39095A | Adverse effect of salicylates, initial encounter |

| | |
|---|---|
| T391X5A | Adverse effect of 4-Aminophenol derivatives, initial encounter |
| T392X5A | Adverse effect of pyrazolone derivatives, initial encounter |
| T39315A | Adverse effect of propionic acid derivatives, initial encounter |
| T39395A | Adverse effect of other nonsteroidal anti-inflammatory drugs [NSAID], initial encounter |
| T394X5A | Adverse effect of antirheumatics, not elsewhere classified, initial encounter |
| T398X5A | Adverse effect of other nonopioid analgesics and antipyretics, NEC, initial encounter |
| T3995XA | Adverse effect of unspecified nonopioid analgesic, antipyretic and antirheumatic, initial encounter |
| T400X5A | Adverse effect of opium, initial encounter |
| T402X5A | Adverse effect of other opioids, initial encounter |
| T403X5A | Adverse effect of methadone, initial encounter |
| T404X5A | Adverse effect of other synthetic narcotics, initial encounter |
| T405X5A | Adverse effect of cocaine, initial encounter |
| T40605A | Adverse effect of unspecified narcotics, initial encounter |
| T40695A | Adverse effect of other narcotics, initial encounter |
| T407X5A | Adverse effect of cannabis (derivatives), initial encounter |
| T40905A | Adverse effect of unspecified psychodysleptics [hallucinogens], initial encounter |
| T40995A | Adverse effect of other psychodysleptics [hallucinogens], initial encounter |
| T415X5A | Adverse effect of therapeutic gasses, initial encounter |
| T420X5A | Adverse effect of hydantoin derivatives, initial encounter |
| T421X5A | Adverse effect of iminostilbenes, initial encounter |
| T422X5A | Adverse effect of succinimides and oxazolidinediones, initial encounter |
| T423X5A | Adverse effect of barbiturates, initial encounter |
| T424X5A | Adverse effect of benzodiazepines, initial encounter |
| T425X5A | Adverse effect of mixed antiepileptics, initial encounter |
| T426X5A | Adverse effect of other antiepileptic and sedative-hypnotic drugs, initial encounter |
| T4275XA | Adverse effect of unspecified antiepileptic and sedative-hypnotic drugs, initial encounter |
| T428X5A | Adverse effect of antiparkinsonism drugs & other central muscle-tone depressants, initial encounter |
| T43015A | Adverse effect of tricyclic antidepressants, initial encounter |
| T43025A | Adverse effect of tetracyclic antidepressants, initial encounter |
| T431X5A | Adverse effect of monoamine-oxidase-inhibitor antidepressants, initial encounter |
| T43205A | Adverse effect of unspecified antidepressants, initial encounter |
| T43215A | Adverse effect of selective serotonin and norepinephrine reuptake inhibitors, initial encounter |
| T43225A | Adverse effect of selective serotonin reuptake inhibitors, initial encounter |
| T43295A | Adverse effect of other antidepressants, initial encounter |
| T433X5A | Adverse effect of phenothiazine antipsychotics and neuroleptics, initial encounter |
| T434X5A | Adverse effect of butyrophenone and thiothixene neuroleptics, initial encounter |
| T43505A | Adverse effect of unspecified antipsychotics and neuroleptics, initial encounter |
| T43595A | Adverse effect of other antipsychotics and neuroleptics, initial encounter |
| T43605A | Adverse effect of unspecified psychostimulants, initial encounter |
| T43615A | Adverse effect of caffeine, initial encounter |
| T43625A | Adverse effect of amphetamines, initial encounter |
| T43635A | Adverse effect of methylphenidate, initial encounter |
| T43695A | Adverse effect of other psychostimulants, initial encounter |
| T438X5A | Adverse effect of other psychotropic drugs, initial encounter |
| T4395XA | Adverse effect of unspecified psychotropic drug, initial encounter |
| T440X5A | Adverse effect of anticholinesterase agents, initial encounter |
| T441X5A | Adverse effect of other parasympathomimetics [cholinergics], initial encounter |
| T442X5A | Adverse effect of ganglionic blocking drugs, initial encounter |
| T442X5S | Adverse effect of ganglionic blocking drugs, sequela |
| T443X5A | Adverse effect of other parasympatholytics and spasmolytics, initial encounter |
| T444X5A | Adverse effect of predominantly alpha-adrenoreceptor agonists, initial encounter |
| T445X5A | Adverse effect of predominantly beta-adrenoreceptor agonists, initial encounter |
| T446X5A | Adverse effect of alpha-adrenoreceptor antagonists, initial encounter |
| T447X5A | Adverse effect of beta-adrenoreceptor antagonists, initial encounter |
| T448X5A | Adverse effect of centrally-acting and adrenergic-neuron-blocking agents, initial encounter |
| T44905A | Adverse effect of unspecified drugs primarily affecting the autonomic nervous system, init. enc. |
| T44905S | Adverse effect of unspecified drugs primarily affecting the autonomic nervous system, sequela |
| T44995A | Adverse effect of other drug primarily affecting the autonomic nervous system, initial encounter |
| T44995S | Adverse effect of other drug primarily affecting the autonomic nervous system, sequela |
| T450X5A | Adverse effect of antiallergic and antiemetic drugs, initial encounter |
| T451X5A | Adverse effect of antineoplastic and immunosuppressive drugs, initial encounter |

| | |
|---|---|
| T452X5A | Adverse effect of vitamins, initial encounter |
| T453X5A | Adverse effect of enzymes, initial encounter |
| T454X5A | Adverse effect of iron and its compounds, initial encounter |
| T45515A | Adverse effect of anticoagulants, initial encounter |
| T45525A | Adverse effect of antithrombotic drugs, initial encounter |
| T45605A | Adverse effect of unspecified fibrinolysis-affecting drugs, initial encounter |
| T45615A | Adverse effect of thrombolytic drugs, initial encounter |
| T45625A | Adverse effect of hemostatic drug, initial encounter |
| T45695A | Adverse effect of other fibrinolysis-affecting drugs, initial encounter |
| T457X5A | Adverse effect of anticoagulant antagonists, vitamin K and other coagulants, initial encounter |
| T458X5A | Adverse effect of other primarily systemic and hematological agents, initial encounter |
| T4595XA | Adverse effect of unspecified primarily systemic and hematological agent, initial encounter |
| T460X5A | Adverse effect of cardiac-stimulant glycosides and drugs of similar action, initial encounter |
| T461X5A | Adverse effect of calcium-channel blockers, initial encounter |
| T462X5A | Adverse effect of other antidysrhythmic drugs, initial encounter |
| T463X5A | Adverse effect of coronary vasodilators, initial encounter |
| T464X5A | Adverse effect of angiotensin-converting-enzyme inhibitors, initial encounter |
| T465X5A | Adverse effect of other antihypertensive drugs, initial encounter |
| T466X5A | Adverse effect of antihyperlipidemic and antiarteriosclerotic drugs, initial encounter |
| T467X5A | Adverse effect of peripheral vasodilators, initial encounter |
| T468X5A | Adverse effect of antivaricose drugs, including sclerosing agents, initial encounter |
| T46905A | Adverse effect of unspecified agents primarily affecting the cardiovascular system, initial encounter |
| T46995A | Adverse effect of other agents primarily affecting the cardiovascular system, initial encounter |
| T470X5A | Adverse effect of histamine H2-receptor blockers, initial encounter |
| T471X5A | Adverse effect of other antacids and anti-gastric-secretion drugs, initial encounter |
| T472X5A | Adverse effect of stimulant laxatives, initial encounter |
| T473X5A | Adverse effect of saline and osmotic laxatives, initial encounter |
| T474X5A | Adverse effect of other laxatives, initial encounter |
| T475X5A | Adverse effect of digestants, initial encounter |
| T476X5A | Adverse effect of antidiarrheal drugs, initial encounter |
| T477X5A | Adverse effect of emetics, initial encounter |
| T478X5A | Adverse effect of other agents primarily affecting gastrointestinal system, initial encounter |
| T4795XA | Adverse effect of unspecified agents primarily affecting the gastrointestinal system, initial encounter |
| T480X5A | Adverse effect of oxytocic drugs, initial encounter |
| T481X5A | Adverse effect of skeletal muscle relaxants [neuromuscular blocking agents], initial encounter |
| T48205A | Adverse effect of unspecified drugs acting on muscles, initial encounter |
| T48295A | Adverse effect of other drugs acting on muscles, initial encounter |
| T483X5A | Adverse effect of antitussives, initial encounter |
| T484X5A | Adverse effect of expectorants, initial encounter |
| T485X5A | Adverse effect of other anti-common-cold drugs, initial encounter |
| T486X5A | Adverse effect of antiasthmatics, initial encounter |
| T48905A | Adverse effect of unspecified agents primarily acting on the respiratory system, initial encounter |
| T48995A | Adverse effect of other agents primarily acting on the respiratory system, initial encounter |
| T490X5A | Adverse effect of local antifungal, anti-infective and anti-inflammatory drugs, initial encounter |
| T491X5A | Adverse effect of antipruritics, initial encounter |
| T492X5A | Adverse effect of local astringents and local detergents, initial encounter |
| T493X5A | Adverse effect of emollients, demulcents and protectants, initial encounter |
| T494X5A | Adverse effect of keratolytics, keratoplastics, & other hair treatment drugs & prep’s, initial encounter |
| T495X5A | Adverse effect of ophthalmological drugs and preparations, initial encounter |
| T496X5A | Adverse effect of otorhinolaryngological drugs and preparations, initial encounter |
| T497X5A | Adverse effect of dental drugs, topically applied, initial encounter |
| T498X5A | Adverse effect of other topical agents, initial encounter |
| T4995XA | Adverse effect of unspecified topical agent, initial encounter |
| T500X5A | Adverse effect of mineralocorticoids and their antagonists, initial encounter |
| T501X5A | Adverse effect of loop [high-ceiling] diuretics, initial encounter |
| T502X5A | Adverse effect of carbonic-anhydrase inhibitors, benzothiadiazides & other diuretics, initial encounter |
| T503X5A | Adverse effect of electrolytic, caloric and water-balance agents, initial encounter |
| T504X5A | Adverse effect of drugs affecting uric acid metabolism, initial encounter |
| T505X5A | Adverse effect of appetite depressants, initial encounter |
| T506X5A | Adverse effect of antidotes and chelating agents, initial encounter |
| T507X5A | Adverse effect of analeptics and opioid receptor antagonists, initial encounter |

| | |
|---|---|
| T508X5A | Adverse effect of diagnostic agents, initial encounter |
| T50905A | Adverse effect of unspecified drugs, medicaments and biological substances, initial encounter |
| T50A15A | Adverse effect of pertussis vaccine, incl. combinations with a pertussis component, initial encounter |
| T50A25A | Adverse effect of mixed bacterial vaccines without a pertussis component, initial encounter |
| T50A95A | Adverse effect of other bacterial vaccines, initial encounter |
| T50B15A | Adverse effect of smallpox vaccines, initial encounter |
| T50B95A | Adverse effect of other viral vaccines, initial encounter |
| T50Z15A | Adverse effect of immunoglobulin, initial encounter |
| T50Z95A | Adverse effect of other vaccines and biological substances, initial encounter |
| T887XXA | Unspecified adverse effect of drug or medicament, initial encounter |

#### S1.3: Medical record abstraction form

See the supplementary data (“anaphylaxis_review_form_template_VUMC.xlsx”) accompanying this manuscript.

#### S1.4: Sampling strata

Table S5: Final sampling strata and sampling weights used at VUMC and KPWA.

| Site | Path | Identifier | Vaccine-proximal | N eligible | N sampled | Sampling Weight |
|---|---|---|---|---|---|---|
| VUMC | 1 | --- | --- | 904 | 165 | 5.479 |
| | 2 | --- | --- | 177 | 44 | 4.022 |
| | 3 | --- | --- | 202 | 45 | 4.489 |
| KPWA | 1 | Even | 0 | 237 | 27 | 8.778 |
| | 1 | Even | 1 | 9 | 9 | 1 |
| | 1 | Odd | 0 | 209 | 17 | 12.294 |
| | 1 | Odd | 1 | 12 | 12 | 1 |
| | 2 | Even | 0 | 189 | 30 | 6.3 |
| | 2 | Even | 1 | 6 | 6 | 1 |
| | 2 | Odd | 0 | 205 | 17 | 12.059 |
| | 2 | Odd | 1 | 5 | 5 | 1 |
| | 3 | Even | 0 | 67 | 9 | 7.444 |
| | 3 | Even | 1 | 3 | 3 | 1 |
| | 3 | Odd | 0 | 81 | 5 | 16.2 |
| | 3 | Odd | 1 | 5 | 5 | 1 |

### S2: Supplemental Results

#### S2.1: UMLS CUIs discovered using AFEP

Table S6: Anaphylaxis-relevant Unified Medical Language System (UMLS) Concept Unique Identifers (CUIs) discovered using the Automated Feature Extraction for Phenotyping (AFEP) method.

| CUI | Concept Name |
|---|---|
| C0000729 | Abdominal cramps |
| C0000737 | Abdominal pain |
| C0001883 | Airways obstruction |
| C0002792 | Anaphylaxis |
| C0002994 | Angioedema |
| C0003467 | Anxiety |
| C0004096 | Asthma |
| C0005658 | Bite wound |
| C0006266 | Bronchospasm |
| C0007203 | Cardiopulmonary resuscitation |
| C0008031 | Chest pain |
| C0009443 | Common cold |
| C0009676 | Confusion |
| C0010200 | Cough |
| C0011991 | Diarrhea |
| C0012833 | Dizziness |
| C0013182 | Drug allergy |
| C0013404 | Dyspnea |
| C0013604 | Edema |
| C0014236 | Endophthalmitis |
| C0014563 | Epinephrine |
| C0015376 | Extravasation |
| C0015663 | Fasting |
| C0016382 | Flushing |
| C0016462 | Food contamination |
| C0016470 | Food allergy |
| C0018790 | Cardiac arrest |
| C0019825 | Hoarseness |
| C0020517 | Hypersensitivity |
| C0020523 | Immediate hypersensitivity |
| C0020649 | Hypotension |
| C0020683 | Hypovolemic shock |
| C0021368 | Inflammation |
| C0021564 | Insect bite NOS |
| C0021925 | Intubation |
| C0021932 | Endotracheal intubation |
| C0022885 | Laboratory test |
| C0023052 | Laryngeal edema |

| | |
|---|---|
| C0024899 | Mastocytosis |
| C0026821 | Muscle cramp |
| C0027497 | Nausea |
| C0027498 | Nausea and vomiting |
| C0027627 | Metastasis |
| C0028778 | Obstruction |
| C0030193 | Pain |
| C0030252 | Palpitations |
| C0033774 | Pruritus |
| C0035273 | Resuscitation |
| C0036974 | Shock |
| C0036980 | Cardiogenic shock |
| C0037090 | Respiratory symptom |
| C0037296 | Skin test |
| C0038340 | Sting |
| C0038450 | Stridor |
| C0038999 | Swelling |
| C0039070 | Syncope |
| C0039231 | Tachycardia |
| C0040533 | Toxic effect of venom |
| C0041657 | Loss of consciousness |
| C0041755 | Adverse drug reaction |
| C0042109 | Urticaria |
| C0042196 | Vaccination |
| C0042420 | Syncope vasovagal |
| C0042963 | Vomiting |
| C0043144 | Wheezing |
| C0079603 | Immunofluorescence |
| C0079840 | Milk allergy |
| C0087111 | Therapeutic procedure |
| C0149783 | Steroid therapy |
| C0151602 | Facial swelling |
| C0151610 | Tongue edema |
| C0155877 | Allergic asthma |
| C0162297 | Respiratory arrest |
| C0199176 | Prophylaxis |
| C0199470 | Mechanical ventilation |
| C0199747 | Allergy test |
| C0202202 | Protein |

| | |
|---|---|
| C0220787 | Endotracheal aspiration |
| C0220870 | Lightheadedness |
| C0221232 | Welts |
| C0231835 | Tachypnea |
| C0231848 | Air hunger |
| C0232070 | Foreign body aspiration |
| C0232292 | Chest tightness |
| C0235710 | Chest discomfort |
| C0236068 | Swelling of tongue |
| C0236071 | Throat constriction |
| C0238614 | Exposure to allergen |
| C0240211 | Lip swelling |
| C0242073 | Pulmonary congestion |
| C0242184 | Hypoxia |
| C0340865 | Anaphylactoid reaction |
| C0344183 | Exercise-induced anaphylaxis |
| C0347950 | Asthmatic attack |
| C0349790 | Exacerbation of asthma |
| C0392707 | Atopy |
| C0413119 | Wasp sting |
| C0413120 | Bee sting |
| C0413234 | Acute allergic reaction |
| C0426576 | Gastrointestinal symptom NOS |
| C0442856 | Hypoperfusion |
| C0476207 | Giddiness |
| C0476273 | Respiratory distress |
| C0521481 | Mucosal edema |
| C0542571 | Face edema |
| C0543467 | Surgical procedure |
| C0546884 | Hypovolemia |
| C0549249 | Depressed level of consciousness |
| C0554804 | Assisted ventilation |
| C0559469 | Egg allergy |
| C0559470 | Peanut allergy |
| C0559546 | Adverse reaction |
| C0577620 | Allergy to nuts |
| C0577628 | Latex allergy |
| C0586407 | Cutaneous symptom |
| C0595862 | Vasodilatation |

| | |
|---|---|
| C0600228 | Cardiopulmonary arrest |
| C0677500 | Stinging |
| C0685898 | Anaphylactic reaction to food |
| C0700184 | Throat irritation |
| C0700198 | Pulmonary aspiration |
| C0740651 | Abdominal symptom |
| C0740852 | Upper airway obstruction |
| C0743747 | Face angioedema |
| C0744425 | Glucocorticoid therapy |
| C0751535 | Cardiac syncope |
| C0850569 | Allergic rash |
| C0854051 | Allergy to sting |
| C0854649 | Anaphylaxis treatment |
| C0856904 | Fish allergy |
| C0857035 | Acute anaphylaxis |
| C0857353 | Hypotensive |
| C0859897 | Vocal cord dysfunction |
| C0877248 | Adverse event |
| C0947961 | Atopic disorders |
| C1096052 | Venomous sting |
| C1145670 | Respiratory failure |
| C1260880 | Rhinorrhea |
| C1260922 | Abnormal breathing |
| C1261392 | Insect bite allergy |
| C1275515 | Venomous bite |
| C1304200 | Lip angioedema |
| C1306577 | Death |
| C1328414 | Blood tryptase |
| C1504322 | Tryptase increased |
| C1504374 | Antihistamine therapy |
| C1527304 | Allergic reaction |
| C1527344 | Dysphonia |
| C1533685 | Injection |
| C1861783 | Median arcuate ligament syndrome |
| C2939065 | Airway edema |
| C3853540 | Aspirin-exacerbated respiratory disease |
| C4047193 | Epinephrine Auto-Injector |
| C4055482 | Airway compromise |
| C4316895 | Anaphylactic shock |

| | |
|---|---|
| C4324659 | Respiratory angioedema |
| C4510560 | Allergy to insect sting |
| C4728126 | Gastrointestinal spasm |
| C5208132 | Respiratory compromise |

#### S2.2: Mapping between MedDRA preferred terms and associated child terms

See the supplementary data ("AFEP_normalization_mapping.xlsx") accompanying this manuscript.

#### S2.3: Supplemental figures

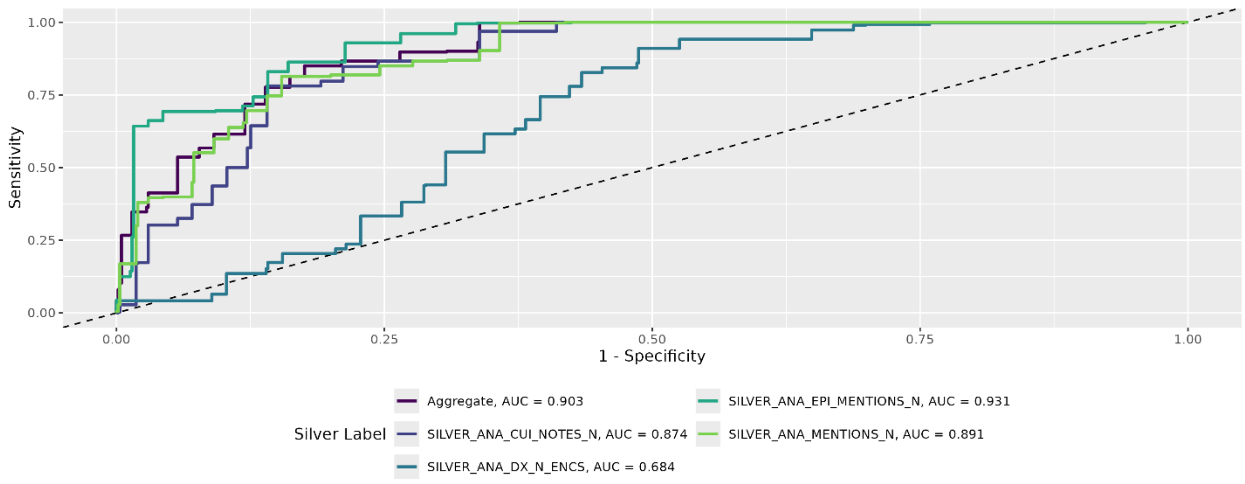


*Figure S1: Receiver operating characteristic curves and AUCs for all models trained at KPWA and evaluated on data from KPWA. The silver labels are counts of anaphylaxis diagnosis codes (ANA_DX_N_ENCS), anaphylaxis mentions (ANA_MENTIONS_N), anaphylaxis concept unique identifiers (ANA_CUI_NOTES_N), and anaphylaxis or epinephrine mentions (ANA_EPI_MENTIONS_N).*

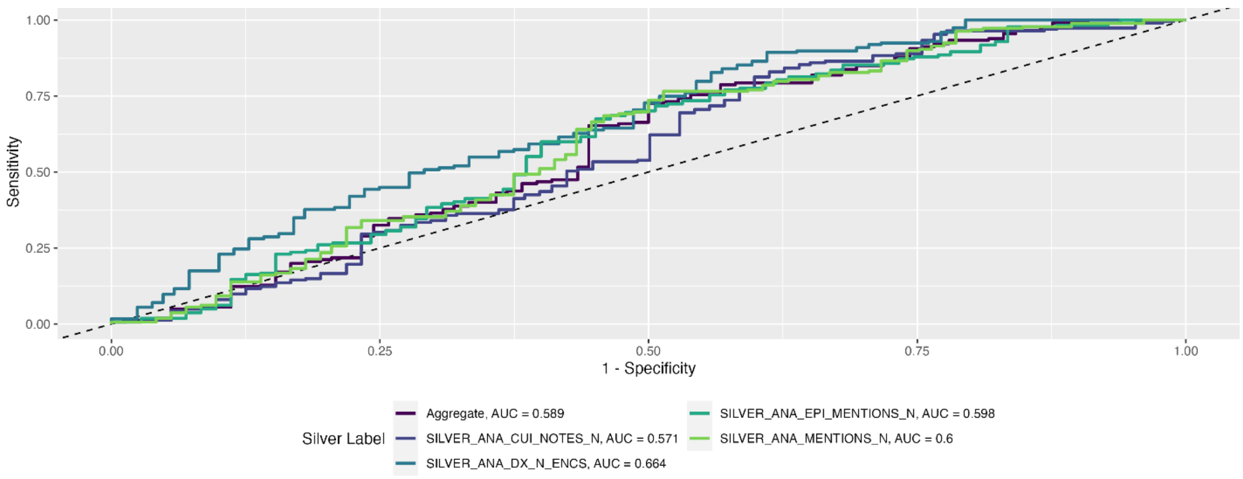


*Figure S2: Receiver operating characteristic curves and AUCs for all models trained at VUMC and evaluated on data from VUMC. The silver labels are counts of anaphylaxis diagnosis codes (ANA_DX_N_ENCS), anaphylaxis mentions (ANA_MENTIONS_N), anaphylaxis concept unique identifiers (ANA_CUI_NOTES_N), and anaphylaxis or epinephrine mentions (ANA_EPI_MENTIONS_N).*

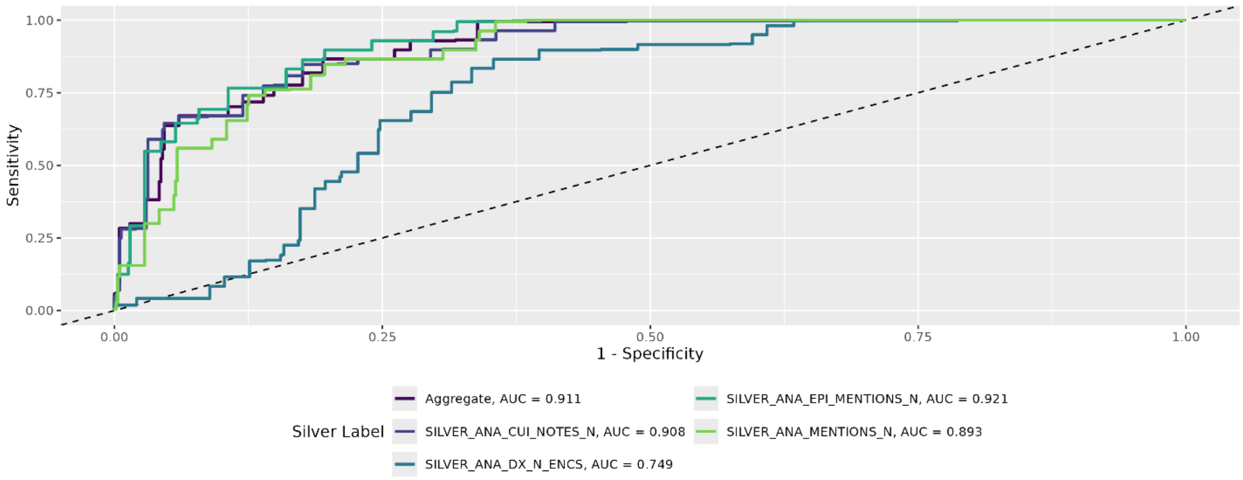


*Figure S3: Receiver operating characteristic curves and AUCs for all models trained at VUMC and evaluated on data from KPWA. The silver labels are counts of anaphylaxis diagnosis codes (ANA_DX_N_ENCS), anaphylaxis mentions (ANA_MENTIONS_N), anaphylaxis concept unique identifiers (ANA_CUI_NOTES_N), and anaphylaxis or epinephrine mentions (ANA_EPI_MENTIONS_N).*

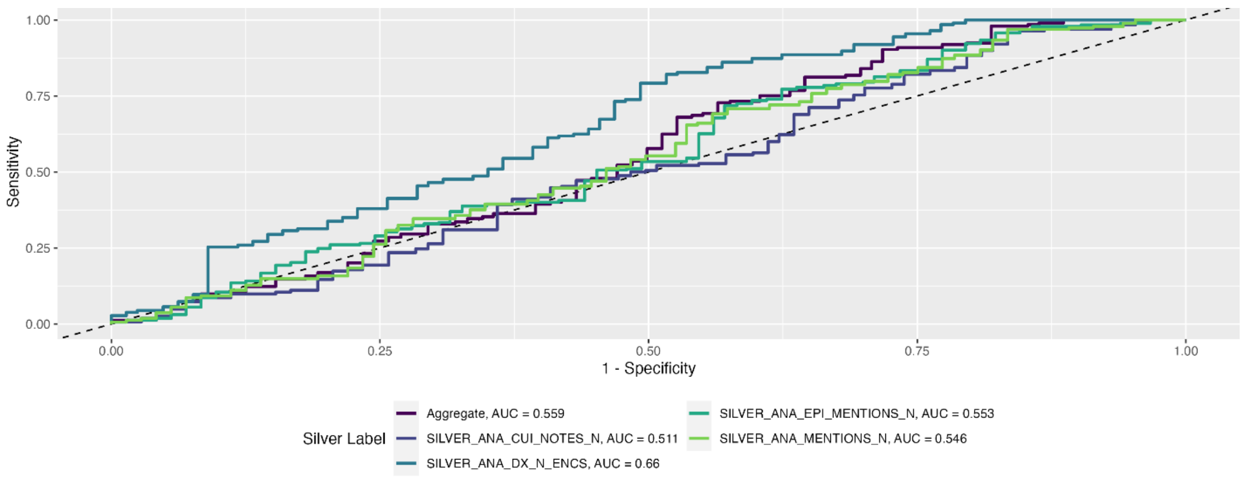


*Figure S4: Receiver operating characteristic curves and AUCs for all models trained at KPWA and evaluated on data from VUMC. The silver labels are counts of anaphylaxis diagnosis codes (ANA_DX_N_ENCS), anaphylaxis mentions (ANA_MENTIONS_N), anaphylaxis concept unique identifiers (ANA_CUI_NOTES_N), and anaphylaxis or epinephrine mentions (ANA_EPI_MENTIONS_N).*
