## Supplemental Section 1.3 Medical record abstraction form instructions for "Identifying anaphylaxis using weakly-supervised prediction models and natural language processing"

### Sentinel Scalable NLP - Anaphylaxis Chart Reviews

#### *Definitions*

Anaphylaxis has multiple accepted definitions…

*A serious life-threatening generalized or systemic hypersensitivity reaction.*

*A serious allergic reaction that is rapid in onset and might cause death.*

*A severe life-threatening generalized or systemic hypersensitivity reaction.*

*A serious allergic reaction that involves more than one organ system (for example, skin, respiratory tract, and/or gastrointestinal tract). It can begin very rapidly, and symptoms may be severe or life-threatening.*

*A severe, life-threatening systemic hypersensitivity reaction characterized by being rapid in onset with potentially life-threatening airway, breathing, or circulatory problems and is usually, although not always, associated with skin and mucosal changes.*

#### *Criteria*

Anaphylaxis determination is based on criteria from the World Health Organization, World Allergy Organization, and National Institute of Alle

The criteria is the source of the questions you will be answering.

We will be basing the final determination on these questions answered by reviewers.

Still, there is a question asking for your opinion on whether you think it is likely anaphylaxis… do not feel too much pressure on this question, calculate whether they meet criteria based on the questions. But we would like to have your opinion for comparison.

#### *Instructions*

For the Group1 sample, you will be reviewing 30 patients each. You will be reviewing the same patients.

Group1 and Group2 will be used to improve the chart review instrument and determine inter-rater ageeement.

You may consult with one another (or other members of the study team) regarding all review groups after Group1 and Group2.

Please let us know if you have any question/concerns/suggestions regarding the questionnaire/form.

The next tab contains the chart review questions in the first/second columns.
Each column represents a patient, and includes StudyID, MRN, and Index Date - the date on which we discovered electronic evidence indicati
Each colored row corresponds to a question.
Descriptions or examples are provided in italics beneath most questions.

You can think of each questions as being prefaced with the phrase, "*Based on the information available to you in the chart…* "
For example, for question **#1**, the question can be read as "*Based on the information available to you in the chart, was there an* **acute onset c**
Answer each question with either **YES** or **NO**.
You may answer with **UNCERTAIN**, but we are hoping most answers will be **YES/NO**.
If you do need to put **UNCERTAIN**, please explain in the optional comments box (last question, **#16**).
If something is not mentioned in the chart, the answer to the question should be **NO**.

Please save the excel file with your name or initials so it can be linked back to the reviewer who completed it (i.e., JCS_anaphylaxis_reviews.G

#### *Additional Guidance*

If the symptom is acute in nature, but potentially related to another disease process - this would be considered a "yes"
However, chronic symptoms known to be the patient's baseline would not qualify. Consider the presence of other symptoms instead.
Feel free to place comments pertaining to specific quesgtions on the lines in-between the YES/NO/Uncertain answers

#### *Questions from Initial Review with Answers*

1. Does a diagnosis of anaphylaxis count for question 12? Or does a provider have to mention it in the note?
    a. Affirmative mention of anaphylaxis should count as a “yes” for this question. A mention of “rule out anaphylaxis” or “suspicion for anap
2. What signs/symptoms count for hypotension? Example: tachycardia or feeling warm
    a. Tachycardia and feeling warm are too generic for an implicit hypotension sign or symptom. We should use the signs/symptoms mentio
3. Does nausea alone count for question 5?
    a. Count any GI symptoms as “Yes” or does it have to be mentioned as “severe/persistent?” Mild nausea or nausea alone would not coun
4. Do we consider any food or medication a likely exposure?
    a. Not for all foods, just common food allergies. All medications and some foods.

5. Do we mark “Yes” for any of the questions related to symptoms even if they are noted to be d/t another disease process other than anaphyla[illegible]
    a. Yes
6. Does mild shortness of breath count for resp compromise?
    a. Yes, count any variation of shortness of breath/dyspnea
7. Would any mention of organ failure count for question #4? Example: AKI or resp failure
    a. Focus on symptoms mentioned on spreadsheet and not just a diagnosis of actual organ failures.
8. What symptoms do we count as bronchospasm? Would chest tightness count?
    a. Only mark specific mentions of bronchospasm.
