## Supplemental Section 1.3 Medical record abstraction form for "Identifying anaphylaxis using weakly-supervised prediction models and natural language processing"

| | | # | 1 | 2 |
|---|---|---|---|---|
| | | study_id | | |
| | | medical record number | | |
| | | index_date | | |
| **1** | **Was there an acute onset of illness?** | | | |
| | *Be generous on interpretting this; it may not specifically say "acute onset"* | | | |
| **2** | **Involvement of the skin and/or mucosal tissue?** | | | |
| | *e.g., generalized hives, pruritus or flushing, swollen lips-tongue-uvula, etc.* | | | |
| **3** | **Respiratory compromise?** | | | |
| | *e.g., dyspnea, wheeze-bronchospasm, stridor, reduced PEF, hypoxemia, etc.* | | | |
| **4** | **Associated symptoms of end-organ dysfunction?** | | | |
| | *e.g., hypotonia (collapse), syncope, incontinence…* | | | |
| **5** | **Severe/persistent gastrointestinal symptoms?** | | | |
| | *e.g., crampy abdominal pain, vomiting* | | | |
| **6** | **Exposure to a *known* allergen *for that patient* ?** | | | |
| | *Known allergen means "patient known to be allergic to it"* | | | |
| **7** | **Exposure to a *likely* allergen?** | | | |

| | |
|---|---|
| *Does not have to be "known" for patient, but is something known to cause alleric reactions in some people* | |
| **8** | **Hypotension, explicit (i.e., documented)** |
| *Documented as (1) hypotension in the text, (2) systolic BP of less than 90 mm Hg, or (3) greater than 30% decrease from that person's baseline* | |
| **9** | **Hypotension, implicit (i.e., associated symptoms)** |
| *Mention of symptoms probable for acute hypotension; e.g., dizziness, lightheadedness, syncope, loss of conciousness, weakness, blurred vision, fatigue* | |
| **10** | **Bronchospasm?** |
| **11** | **Laryngeal involvement?** |
| *e.g., stridor, vocal changes, odynophagia, etc.* | |
| **12** | **Is there an explicit diagosis of anaphylaxis?** |
| *Provider has documented in words that there is likely anaphylaxis; this is not referring to any diagnosis code* | |
| **13** | **Does the preponderance of evidence suggest this was a case of anaphylaxis?** |
| *This is based on your own opinion* | |
| **14** | **Was this review difficult or complex?** |

| | |
|---|---|
| *e.g., did you have a hard time finding relevant information? Was the diagnosis complicated in some way?* | |
| **15** | **Are you uncertain about this case?** |
| *e.g., you cannot make a determination; you wish to consult other reviewers, etc.* | |
| **16** | **Optional comments** |
